# Data-driven neuroanatomical subtypes of primary progressive aphasia

**DOI:** 10.1101/2024.05.13.24307283

**Authors:** Beatrice Taylor, Martina Bocchetta, Cameron Shand, Emily G Todd, Anthipa Chokesuwattanaskul, Sebastian J Crutch, Jason D Warren, Chris JD Hardy, Jonathan D Rohrer, Neil P Oxtoby

## Abstract

The primary progressive aphasias are rare, language-led dementias, with three main variants: semantic, non-fluent/agrammatic, and logopenic. Whilst semantic variant has a clear neuroanatomical profile, the non-fluent/agrammatic and logopenic variants are difficult to discriminate from neuroimaging. Previous phenotype-driven studies have characterised neuroanatomical profiles of each variant on MRI. In this work we used a machine learning algorithm known as SuStaIn to discover data-driven neuroanatomical “subtype” progression profiles and performed an in-depth subtype–phenotype analysis to characterise the heterogeneity of primary progressive aphasia.

Our study included 270 participants with primary progressive aphasia seen for research in the UCL Queen Square Institute of Neurology Dementia Research Centre, with follow-up scans available for 137 participants. This dataset included individuals diagnosed with all three main variants (semantic: n=94, non-fluent/agrammatic: n=109, logopenic: n=51) as well as individuals with un-specified primary progressive aphasia (n=16). A data set of 66 patients (semantic n=37, non-fluent/agrammatic: n=29) from the ALLFTD North American cohort study, was used to validate our results. MRI scans were segmented and SuStaIn was employed on 19 regions of interest to identify neuroanatomical profiles independent of the diagnosis. We assessed the assignment of subtypes and stages, as well as their longitudinal consistency.

We discovered four neuroanatomical subtypes of primary progressive aphasia, labelled S1, S2, S3, S4, exhibiting robustness to statistical scrutiny. S1 correlated strongly with semantic variant, while S2, S3, and S4 showed mixed associations with the logopenic and non-fluent/agrammatic variants. Notably, S3 displayed a neuroanatomical signature akin to a logopenic only signature, yet a significant proportion of logopenic cases were allocated to S2. The non-fluent/agrammatic variant demonstrated diverse associations with S2, S3, and S4. No clear relationship emerged between any of the neuroanatomical subtypes and the unspecified cases. At first follow up 84% of patients subtype assignment was stable, and 91.9% of patients stage assignment was stable. The ALLFTD dataset validated the findings.

Our study, leveraging machine learning on a large primary progressive aphasia dataset, delineated four distinct neuroanatomical patterns. The identification of multiple profiles within the logopenic and non-fluent/agrammatic variants, alongside intra-phenotypic overlap, supports recent conceptualisations of primary progressive aphasia as a spectrum of disorders rather than discrete entities. Understanding the multifaceted profiles of the disease, encompassing neuroanatomical, molecular, clinical, and cognitive dimensions, holds potential implications for clinical decision support.

## Introduction

Primary progressive aphasia (PPA) is a term used to refer to a set of rare, language-led dementias. There are three canonical variants of PPA recognised by consensus diagnostic criteria: semantic (svPPA), nonfluent/agrammatic (nfvPPA), and logopenic (lvPPA). The classification of PPA into one of the three major variants can occur at one of three levels: clinical, imaging-supported, or definite pathological. At the clinical level, svPPA is associated with impaired naming and single-word comprehension as the initial manifestations of a pan-modal semantic memory impairment^1^. The clinical spectrum of nfvPPA is the most diverse of the three canonical variants and is typically associated with agrammatism and/or apraxia of speech.^1–3^ The most recently defined of the PPA variants is lvPPA, having been first described in 2004^4^ and enshrined in consensus criteria in 2011.^5^ It is typified by impaired word retrieval, and impaired repetition of phrases.^1,6^ The term PPA also encompasses further ‘fragmentary’ syndromes,^1,6^ including mixed or atypical presentations.^7–9^ In a research setting, individuals who do not fit the profile of one of the three main variants but who satisfy overarching criteria for PPA (i.e. a progressive, speech/language-led dementia) may be classified as PPA not otherwise specified (PPA-nos).^8^ As the disorder progresses people with all PPA variants may experience non-language cognitive impairments including memory difficulties, executive function deficits, and behavioural changes.^10^

Neuroimaging may be used to corroborate a diagnosis based on symptom presentation^1,11,12^. The clearest anatomical progression occurs in svPPA, which is typically associated with left anterior temporal lobe atrophy. Recent studies have identified the initial phase of atrophy in svPPA as beginning in the bilateral amygdala and progressing through the left, then right temporal pole.^13,14^ Neuroimaging findings in nfvPPA, lvPPA and PPA-nos are more variable,^6^ with neuroanatomical similarities between nfvPPA and lvPPA resulting in difficulties discriminating these phenotypes based on MRI scans alone.^2,15^ A recent study found an association between a clinical diagnosis of nfvPPA and initial atrophy of the left striatum, followed by the bilateral anterior insula, then the left thalamus with relative sparing of the left precuneus and cuneus.^14^ A study of the neuroanatomical profile of lvPPA, identified initial atrophy of the left posterior temporal lobe, with longitudinal involvement of the parietal lobe.^16^ Whether PPA-nos cases share a common underlying neuroanatomical profile is unknown.^9^ There is therefore considerable clinical and research interest in defining the neuroanatomical underpinnings of PPA.

A clinical diagnosis of PPA is typically associated with a primary pathology of frontotemporal lobar degeneration (FTLD) or Alzheimer’s disease (AD).^17,18^ There is pathological variation by clinical variant: over 80% of svPPA diagnoses are found to be caused by the TAR DNA-binding protein 43 (FTLD-TDP43) proteinopathy at post-mortem^19^; over 80% of nfvPPA diagnoses are associated with a primary tauopathy (FTLD-tau)^19^; and over 80% of lvPPA diagnoses are associated with Alzheimer’s disease pathology.^20^ Genetic cases, typically caused by mutations in progranulin (*GRN*), or more rarely chromosome 9 open reading frame 72 (*C9orf72*), represent a substantial minority of all PPA cases. Genetic PPA has been reported as most predominant in nfvPPA and PPA-nos.^21^

The overlap of clinical phenotype and neuroanatomical changes in PPA is poorly understood. Previous studies have investigated neuroanatomy within PPA phenotypes,^14,22^ which we sought to advance by providing (a) a probabilistic characterisation of neuroanatomy within clinical phenotype, and (b) a data-driven discovery of neuroanatomical subtypes of PPA. We applied the SuStaIn algorithm^23^ to a large retrospective dataset of MRI scans from individuals diagnosed with PPA. We analysed model stability in a longitudinal subset of participants with PPA and validated the neuroanatomical subtypes using an external dataset. Finally, we investigated the correspondence between model subtypes and clinical phenotypes to better understand neuroanatomical heterogeneity amongst the canonical variants of PPA. These advancements (a) provide a quantitative template for understanding disease progression and for neuroanatomical assessment/staging of patients, potentially before symptoms appear; and (b) contribute to our understanding of biological overlap between clinical variants.

## Materials and methods

We performed discovery on a historical dataset from the UCL Queen Square Institute of Neurology Dementia Research Centre in London UK, and used data from the North American ALLFTD study as an external test set.

### Sample Characteristics

Data were collected from participants enrolled in five longitudinal studies at the Dementia Research Centre, UCL, between 1993 and 2020, further details of which are provided in Supplementary Material. All participants gave informed consent, and ethical approval was granted by The National Hospital for Neurology and Neurosurgery & UCL Institute of Neurology Joint Research Ethics Committee. The Queen Square discovery dataset was compiled retrospectively from these studies, with inclusion criteria of a PPA diagnosis according to clinical consensus criteria,^1^ and an MRI scan. The dataset included 270 patients with a diagnosis of PPA (svPPA n=94; nfvPPA n=109; lvPPA n=51; PPA-nos n=16). Of those with svPPA, n=89 were ‘left predominant’, classified according to whether the left or right temporal pole was more atrophied at baseline.^24,25^ The 53% of cases that were diagnosed prior to 2011 (before the publication of the current diagnostic criteria) were retrospectively assessed by a senior neurologist (JDR), and re-diagnosed/re-labelled where appropriate. A minority of patients received a secondary clinical diagnosis of an associated neurological condition: Parkinson’s disease (PD) (n=1), progressive supranuclear palsy (PSP) (n=8), corticobasal syndrome (CBS) (n=8), PSP/CBS (n=1) and motor neurone disease (MND) (n=2).

Histopathological data was available for 51 (18%) patients in the cohort, whose brains were donated to the Queen Square Brain Bank archives, where tissues are stored under a licence from the Human Tissue authority (No. 12198). Both the brain donation programme and protocols have received ethical approval for donation and research by the NRES Committee London—Central. Among these cases, 14 presented with Alzheimer’s disease pathology, including one case with concomitant Parkinson’s disease (AD-PD). Fourteen cases exhibited FTLD-tau pathology, with eight diagnosed as Pick’s disease (Tau-Pick’s), four as corticobasal degeneration (Tau-CBD), two as globular glial tauopathy (Tau-GGT), and two as progressive supranuclear palsy (Tau-PSP). Additionally, 21 cases presented with FTLD-TDP-43 pathology, with one categorised as Type A and 20 as Type C.

Follow-up scans were obtained from 137 (of 270) participants during subsequent research visits. The interval between consecutive scans was 1.1 ± 0.6 years (mean ± standard deviation). It is important to note that we included follow-up scans even if the MRI scanner was different to those used at baseline; further details are included in the Supplementary material.

For the w-scoring, we used MRI scans from 121 cognitively normal control subjects, recruited from various studies at the Dementia Research Centre, UCL.

Our test dataset was formed of 66 members of ALLFTD (www.allftd.org/), a large North American natural history study of FTD, who had a diagnosis of PPA and an MRI scan. This included individuals with svPPA (n=37) and nfvPPA (n=29). There were no individuals with lvPPA in the cohort as inclusion criteria for ALLFTD is assumed frontotemporal lobal degeneration. At least one follow up scan was obtained for every individual in the dataset. For w-scoring of the test dataset we used MRI scans from cognitively normal control subjects (n=121) recruited from the same ALLFTD study. Table 1 presents the demographic characteristics of the cohort.

**Table 1:**
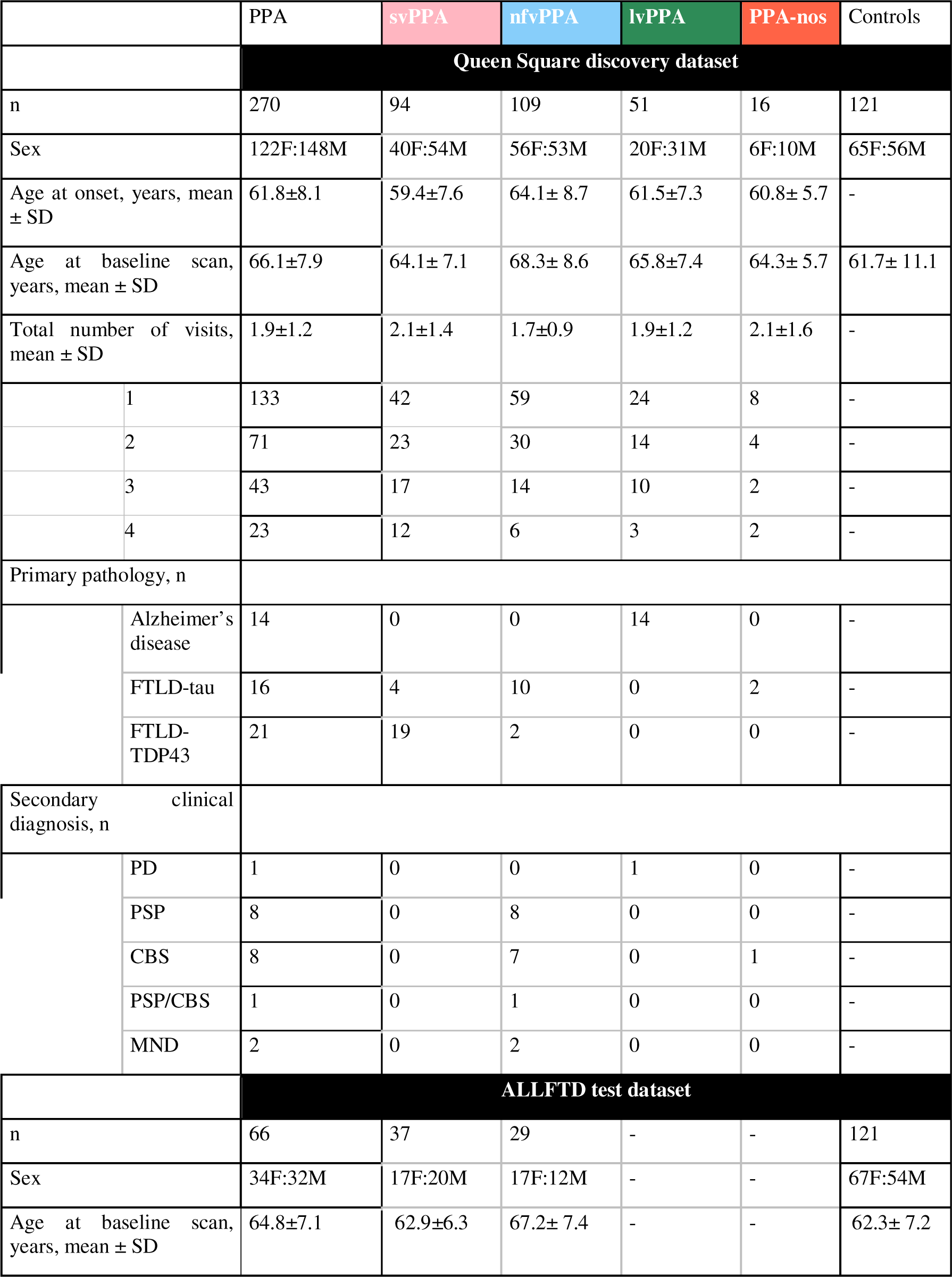
Cohort demographics. Colours are used to distinguish the three phenotypes: svPPA (pale pink), nfvPPA (pale blue), lvPPA (dark green). Abbreviations: PPA – Primary Progressive Aphasia, svPPA – semantic variant PPA, nfvPPA – nonfluent/agrammatic variant PPA, lvPPA – logopenic variant PPA, PPA-nos – PPA not otherwise specified, FTLD-tau – Frontotemporal lobal degeneration tau, FTLD-TDP43 – frontotemporal lobar degeneration TAR DNA-binding protein 43, PD – Parkinson’s disease, PSP – progressive supranuclear palsy, CBS – corticobasal syndrome, MND – motor neurone disease.

### Image acquisition and pre-processing

Participants in the Queen Square discovery dataset underwent MRI scans on various 1.5T or 3T MRI scanners from two different manufacturers (Siemens Trio 3T, Siemens Prisma 3T, GE Signa 1.5T, see Supplementary Table 1). Participants in the ALLFTD dataset underwent MRI scans on various 3T MRI scanners from two different manufacturers (Siemens Trio 3T, GE Signa 3T, GE Discovery 3T, see Supplementary Table 2). In both datasets, cortical and subcortical volumes were parcellated on volumetric T1-weighted MRIs using the geodesic information flows tool (GIF),^26^ which utilises the Neuromorphometrics brain parcellation and is based on atlas propagation and label fusion. The total intracranial volume was calculated using SPM12 v6255, running under Matlab R2014b (MathWorks, Natick, MA).^27^ All segmentations were visually inspected to ensure accurate segmentation. Only subjects with a usable T1-weighted MRI were included. Brain pathology not related to PPA (e.g., brain lesions such as tumours) was an exclusion criterion.

### Statistical analysis

#### Subtype and Stage Inference Algorithm

SuStaIn is an unsupervised machine learning algorithm that probabilistically identifies disease progression subtypes, i.e., it jointly estimates clusters of distinct biomarker trajectories and the trajectories themselves.^23^ These trajectories are characterised as cumulative sequences of biomarkers transitioning from normal to abnormal states, in reference to a control group. In this work said biomarkers are volumes of segmented brain regions, we refer to the group level volumetric abnormalities as atrophy. The versatility of SuStaIn has been demonstrated across various neurodegenerative diseases datasets, effectively modelling tau PET heterogeneity in typical Alzheimer’s disease,^28^ volumetric abnormalities in typical Alzheimer’s disease^23^ and FTD,^29^ and neuroanatomical subtypes in multiple sclerosis.^30^ For our analysis, we employed the Python implementation of the SuStaIn algorithm.^31^

At each patient visit, the w-score (a.k.a., covariate-adjusted z-score) of brain volume from controls was computed as follows:

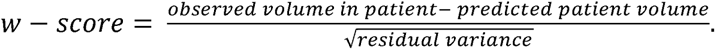

To calculate the predicted patient volume and residual variance, a linear regression was performed on the controls while accounting for covariates of sex, age at scan, total intracranial volume, and scanner types.^13,32^

In the model, biomarker abnormality is determined by reaching user-defined w-score thresholds in the respective region. The number of stages in the identified disease trajectories is equal to the total number of abnormality scores across all biomarkers included in the model. This constrains the number of input biomarkers — as to prevent overfitting the total number of stages across subtypes should not exceed the dataset size.

The selection of regions of interest (ROIs) followed the framework outlined by Scotton *et al.*.^33^ After consultation with neurologists and PPA and neuroimaging experts (MB, JDW and CJDH) we decided to include regions which a) had previously been implicated in PPA^34–36^; and b) were large enough for reliable segmentation. To streamline the model input and reduce feature complexity, some contiguous sub-regions were combined into composite regions where this was appropriate anatomically. This resulted in a final list of 19 ROIs (Supplementary Table 3).

For each of the 19 ROIs we used three w-score thresholds (1,2,3), resulting in 57 model events. We justified the choice of three severity scores since across all the 19 ROIs >5% of the patients had reached the most severe score at baseline, and thus contained sufficient ‘disease signal’ (Supplementary Fig. 2). The model was fit on baseline data, with model comparison under 10-fold cross validation (cross validation information criterion of up to 5 subtypes) used to determine the number of subtypes hyperparameter.^23^ Model uncertainty was estimated using 10,000 MCMC iterations.

#### Similarity measure for data-driven subtypes

We supplemented our model comparison analysis with assessment of subtype statistical similarity to inform model parsimony. We assessed subtype similarity, by comparing posterior event distributions using Hellinger distance.^37^ For two discrete probability distributions *p* = (*p*_1_, … , *p_k_*), and Q = (*q*_1_, … , *q_k_*), the Hellinger distance is defined as:

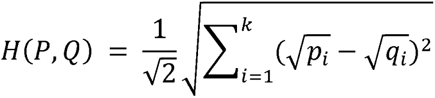

The Hellinger distance between two identical distributions is equal to zero; whilst the maximal Hellinger distance of one occurs when *P* assigns positive probability everywhere *Q* The Hellinger distance between two identical distributions is equal to zero; whilst the assigns zero probability, or vice versa. We calculated a reference value of *H_o_* = 0.89 ± 0.019 (95% CI [0.893, 0.895]) which represents the statistical similarity of randomised models (for further details see the Supplementary material).

#### Subtype characterisation

We used the *X*^2^ contingency test to compare proportions, e.g., when exploring subtype-phenotype relationships.

#### Patient subtyping and staging

SuStaIn provides a patient subtyping and staging mechanism. Patients were assigned to a stage by maximising their stage likelihood across subtypes.^38,39^ Individuals assigned to stage zero were deemed to not be ‘subtypable’ since their regional brain volumes were comparable to controls. Those deemed subtypable were assigned to their most probable subtype.^23^

We visualised the subtypes of neuroanatomical trajectory using positional variance diagrams, which show the posterior distribution of events. ^39^ Blocks of colour represent the sequence (numbered by the stages on the x-axis) in which regional brain volumes (listed on the y-axis) become abnormal. Colours correspond to the severity score threshold reached at that stage, w-score 1 (red), w-score 2 (magenta), w-score 3 (blue). Colour density corresponds to model (un)certainty, with dark colours representing high certainty regarding the position, and lighter colours representing low certainty. We visualised the atrophy pattern of each subtype using BrainPainter.^40^

#### Single phenotype staging

The original dataset was divided based on clinical diagnosis, creating three smaller datasets representing single phenotypes: svPPA, nfvPPA, and lvPPA. This data was used to analyse correspondence between the data-driven subtypes and the expected sequence of disease progression in the clinical variants. For each single phenotype dataset we fit a single Z Score progression model^23^ by using the pySuStaIn software^31^ with N_subtypes = 1 and all other parameters identical.

#### Changing diagnostic criteria consistency

To assess the impact of the 2011 change in diagnostic criteria to include lvPPA as a clinical phenotype^1,5^, we conducted a separate analysis on the set of individuals who received their scan in or after 2011. The full details of this analysis can be found in the Supplementary material.

#### Longitudinal model consistency

The trained model was used to assign longitudinal subtype and stage to patients using available follow-up data. To assess the impact of changing scanner type on our results, we conducted a separate longitudinal analysis on the restricted subset of individuals with longitudinal scans acquired using the same MRI machine as at baseline – the full details of this analysis can be found in the Supplementary material.

#### Test set: ALLFTD

Patients in the ALLFTD cohort were w-scored with respect to controls from the same study, subtypes and stages were assigned to them using the model trained on the Queen Square discovery dataset.

### Data availability

The Queen Square data set that supports the findings of this study are available from the corresponding author, upon reasonable request. The ALLFTD data set is available upon reasonable request from https://www.allftd.org/data.

## Results

### Data-driven neuroanatomical subtypes and stages

Figure 1 shows the four data-driven subtypes of PPA discovered by SuStaIn, supported by cross-validation (Supplementary Fig 3). Figure 2 shows a visualisation of these four subtypes on the brain surface. The identification of four subtypes was supported by statistical pairwise analysis between subtypes, which found low similarity between the positional variance diagrams (Supplementary Table 4).

**Figure 1:**
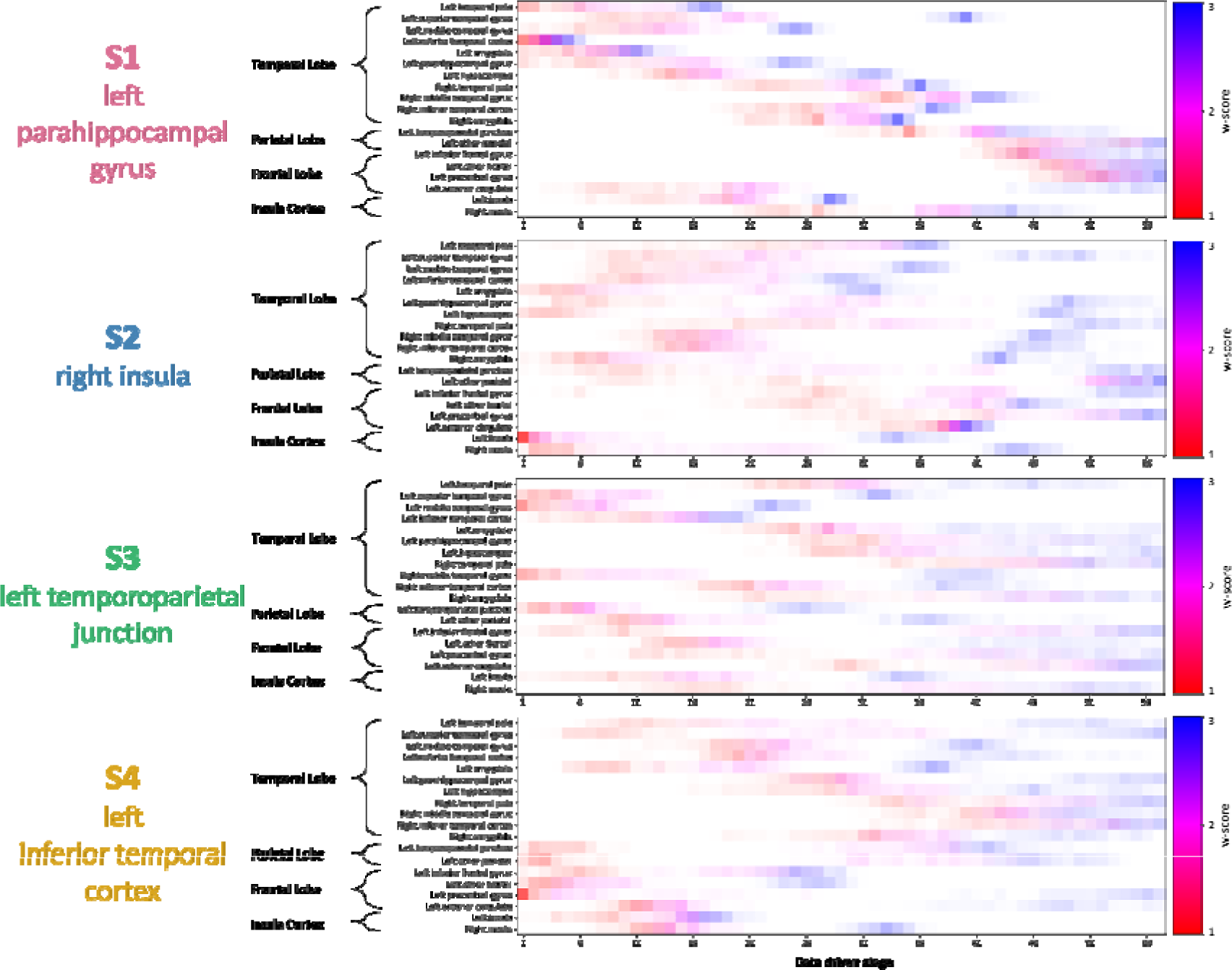
Positional variance diagrams for the data-driven subtypes. Along the y-axis are the regions of interest used in the model, grouped by location in the brain. The data-driven stages correspond to the sequence that brain regions become abnormal, with colour representing degree of abnormality (w-score 1: red, w-score 2: pink, w-score 3: blue), and colour density representing model certainty.

**Figure 2:**
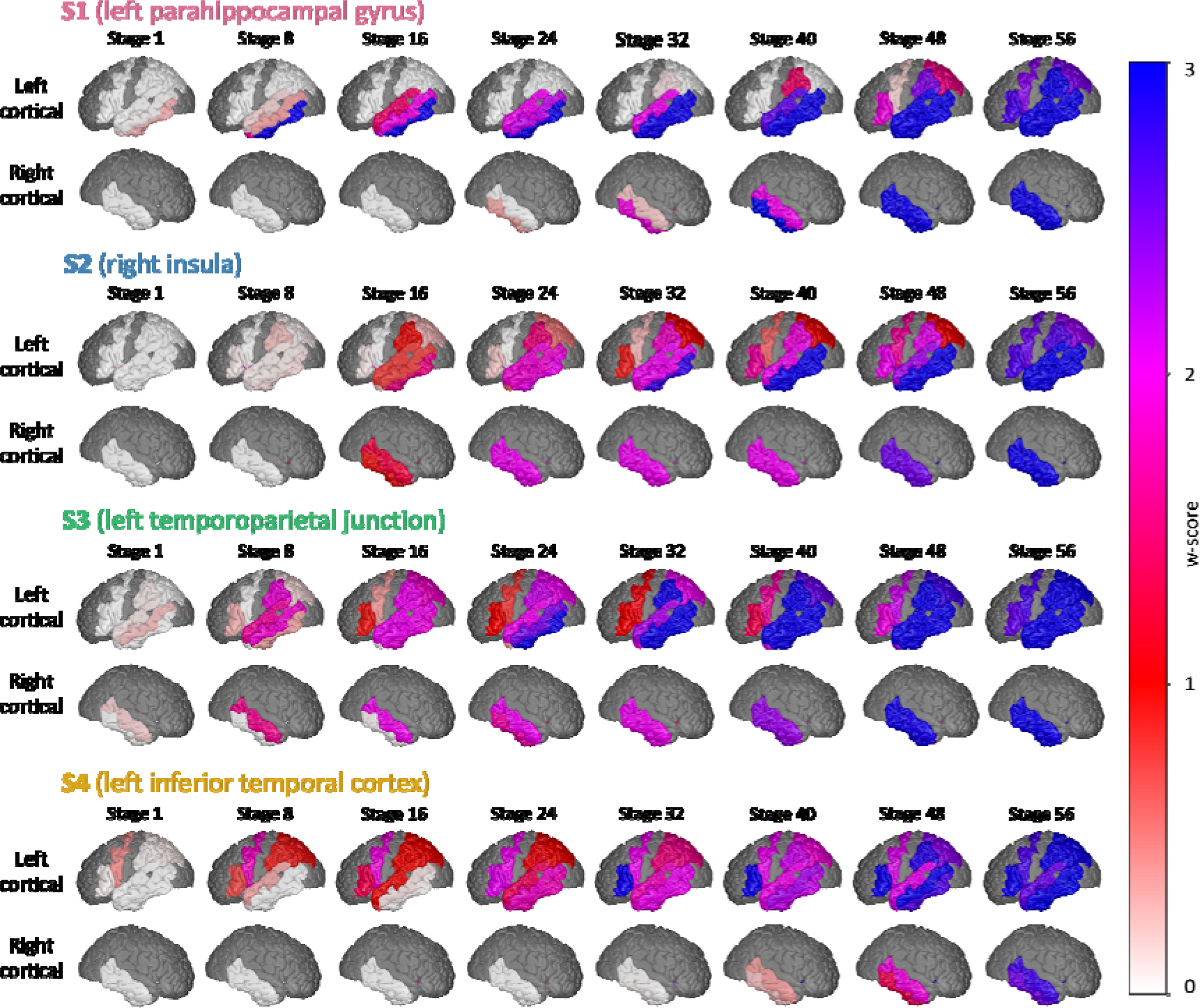
Stages of regional atrophy in the outer cortical left hemisphere for the data-driven subtypes. The colour representing degree of abnormality (w-score 1: red, w-score 2: pink, w-score 3: blue). Brain regions which are dark grey were not included in the model.

The first subtype (S1 – left parahippocampal gyrus) was distinguished by initial atrophy of the left temporal pole, starting in the left inferior temporal cortex which rapidly progressed through all three w-scores. This was followed by atrophy of the insula cortex, then the right temporal pole. There was relative sparing of the left frontal lobe.

In the second subtype (S2 – right insula) there was early atrophy in the left insula followed by the right insula. This was followed by the left hippocampus, the left parahippocampal gyrus and the bilateral amygdala. There was relative sparing of the temporal pole bilaterally. The left anterior cingulate was spared until later in the disease, when it progressed through all w-scores over fewer than ten model stages, suggesting rapid neurodegeneration.

In the third subtype (S3 – left temporoparietal junction) there was initial atrophy in regions around the left temporoparietal junction and the temporal pole. There was relative sparing of the left hippocampus, left amygdala and left parahippocampal gyrus which did not reach the first w-score until a middle stage. Visually compared to S1 and S2, S3 had higher positional variance, due to either lower sample size or simultaneous events.^41^

The fourth, and final subtype, (S4 – left inferior temporal cortex) presented with early atrophy of the left precentral gyrus. This was followed by atrophy of the other frontal lobe regions, then atrophy of the insula cortex. There was relative sparing of the temporal pole bilaterally. Similarly to S3, S4 had higher positional variance.

Across the subtypes, there was a good representation of patients in early and middle stages of the disease, with no-one assigned to a stage above stage 50 of 57 at baseline (Supplementary Fig. 4). In total six individuals were assigned to data-driven stage zero at baseline, and were excluded from all subsequent baseline analysis since they were deemed not ‘subtypable’.

Table 2 records the subtype demographics. Of those who were subtypable, 84% were assigned to a baseline subtype with probability >0.75 (Supplementary Fig. 12). Subtype 1 was the largest subtype, with n=82 (31.0%) patients were assigned to it, with an age at first scan 63.8±6.9 years. Amongst patients assigned to S1, >90% were assigned a disease stage above 15, suggesting most patients assigned to this subtype were beyond the initial disease stages at baseline. Seventy-two (26.9%) patients were assigned S2, with an age at first scan of 66.5±7.2 years; n=59 (22.3%) patients were assigned to S3, with an age at first scan of 66.6±8.3 years; n=52 (19.7%) patients were assigned to S4, with an age at first scan of 68.4±9.2 years.

**Table 2:** Subtype demographics. Colours are used to distinguish the four data driven subtypes: S1 (pink), S2 (blue), S3 (green), S4 (yellow). Abbreviations: PPA – Primary Progressive Aphasia, svPPA – semantic variant PPA, nfvPPA – nonfluent/agrammatic variant PPA, lvPPA – logopenic variant PPA, PPA-nos – PPA not otherwise specified, FTLD-tau – Frontotemporal lobal degeneration tau, FTLD-TDP43 – frontotemporal lobar degeneration TAR DNA-binding protein 43, PD – Parkinson’s disease, PSP – progressive supranuclear palsy, CBS – corticobasal syndrome, MND – motor neurone disease.

|  |  | S1 (left parahippocampal gyrus) | S2 (right insula) | S3 (left temporoparietal junction) | S4 (left inferior temporal cortex) |
| --- | --- | --- | --- | --- | --- |
|  |  | Queen Square discovery dataset |  |  |  |
| n |  | 82 | 71 | 59 | 52 |
| Sex |  | 36F:46M | 34F:37M | 22F:37M | 27F:25M |
| Age at onset, years, mean ± SD | | 59.6±7.5 | 61.8±7.6 | 62.1±8.0 | 64.4±9.0 |
| Age at baseline scan, years, mean ± SD | | 63.8±6.9 | 66.5±7.2 | 66.6±8.3 | 68.4±9.2 |
| Diagnosis, n |  |  |  |  |  |
|  | svPPA | 71 | 15 | 4 | 4 |
|  | nfvPPA | 3 | 34 | 21 | 45 |
|  | lvPPA | 1 | 18 | 31 | 1 |
|  | PPA-nos | 7 | 4 | 3 | 2 |
| Primary pathology, n |  |  |  |  |  |
|  | Alzheimer's disease | 1 | 2 | 11 | 0 |
|  | FTLD-tau | 4 | 4 | 1 | 7 |
|  | FTLD-TDP43 type C | 16 | 3 | 1 | 1 |
| Secondary diagnosis, n |  |  |  |  |  |
|  | PD | 0 | 0 | 1 | 0 |
|  | PSP | 0 | 2 | 1 | 2 |
|  | CBS | 1 | 4 | 1 | 2 |
|  | PSP/CBS | 0 | 0 | 1 | 0 |
|  | MND | 0 | 1 | 1 | 0 |
|  |  | ALLFTD test dataset |  |  |  |
| n |  | 23 | 15 | 9 | 14 |
| Sex |  | 11F:12M | 5F:10M | 4F:5M | 9F:5M |
| Age at baseline scan, years, mean ± SD | | 63.1±6.2 | 63.2±6.3 | 68.6±6.4 | 66.6±8.1 |
| Diagnosis, n |  |  |  |  |  |
|  | svPPA | 20 | 10 | 0 | 7 |
|  | nfvPPA | 3 | 5 | 9 | 7 |

### Association with clinical variant and pathology

Figure 3 shows the association between each participant’s clinical diagnosis and the subtype they were assigned to. The strongest phenotype–subtype concordance occurred for svPPA which made up 86.6% of participants in S1 (left parahippocampal gyrus), and for nfvPPA proportion of clinical diagnoses in each subtype was statistically significant [*X*^2^ (9, *N* = 264) which made up 86.6% of participants assigned to S4 (left inferior temporal cortex). The = 269.4, *p*<.001].

**Figure 3:**
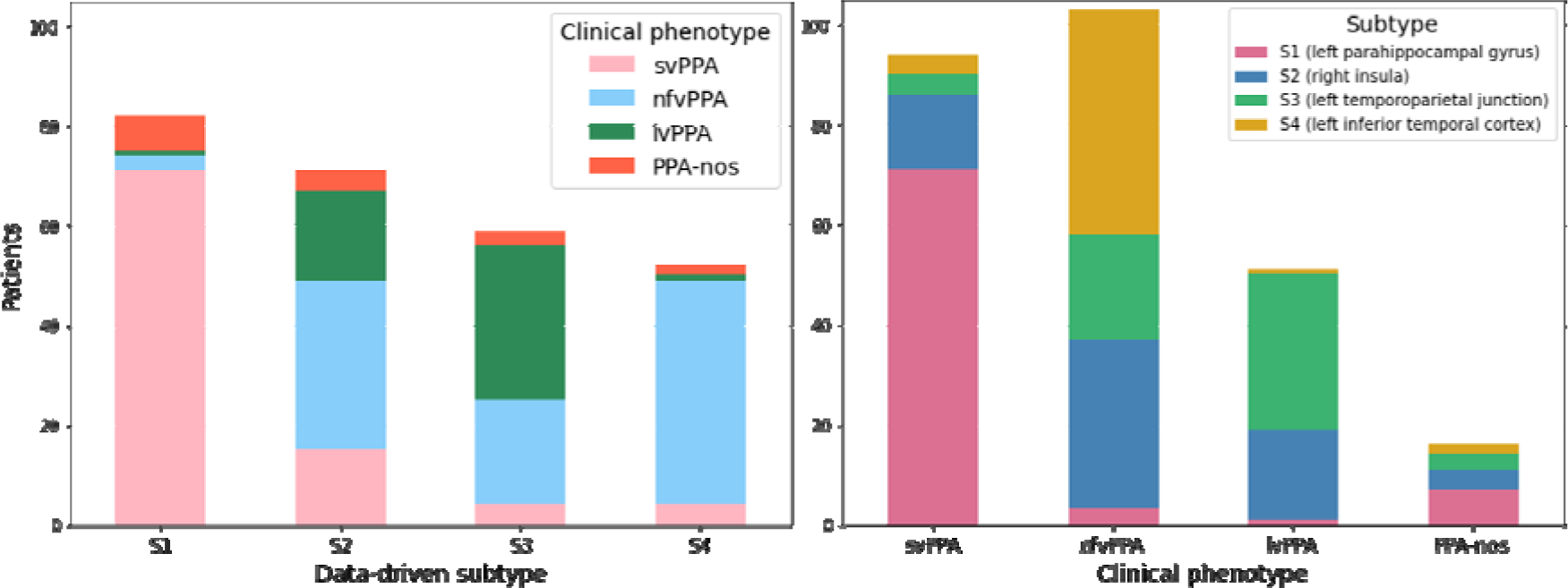
Comparison between data-driven subtype assignment and clinical diagnosis. The stacked bar chart on the left shows the number of patients with each clinical diagnosis by data-driven subtype assignment. The figure on the right shows the number of patients who were assigned to each data-driven subtype by clinical diagnosis. Abbreviations: svPPA – semantic variant PPA, nfvPPA – nonfluent/agrammatic variant PPA, lvPPA – logopenic variant PPA, PPA-nos – PPA not otherwise specified.

The strongest pathology-subtype concordance occurred for FTLD-TDP43 type C for which 16 of 20 (80%) were assigned to S1 (left parahippocampal gyrus), and AD for which 10 of 13 (76.9%) were assigned to S3 (left temporoparietal junction). Further details are in the Supplementary material.

### Comparison with single phenotype staging

Supplementary Fig. 8 shows the positional variance diagrams for the three clinical phenotypes. A diagnosis of svPPA was associated with early decline of the left inferior temporal cortex, followed by decline across the other left temporal regions, then the insula cortex, then the right temporal regions, and finally decline of the frontal regions. A diagnosis of nfvPPA was associated with initial decline of the left precentral gyrus, then the other left frontal lobe regions and the parietal lobe, then the insula cortex, closely followed by the temporal lobe. A diagnosis of lvPPA was associated with initial decline of the left middle temporal gyrus, followed by the superior temporal gyrus, left inferior temporal cortex and left temporoparietal junction, with late sparing of the right temporal pole and the left anterior cingulate.

There was a statistically significant correlation between the neuroanatomical progression in S1 (left parahippocampal gyrus) and the svPPA phenotypic cohort (Hellinger distance: 0.44), and S3 (left temporoparietal junction) and the neuroanatomical progression in the lvPPA phenotypic cohort (Hellinger distance: 0.46) (Supplementary Table 5 and Supplementary Fig. 10).

The analysis of the subset of individuals who were scanned after 2010, and thus after lvPPA was enshrined in the consensus criteria, was consistent with the findings in the main Queen Square discovery dataset – see Supplementary Material.

### Longitudinal data-driven subtypes and staging

Figure 4 is a Sankey diagram demonstrating the longitudinal assignment to subtype for the 135/137 patients who had longitudinal data and were subtypable at baseline. Subtype consistency between first and second MRI scan was: 87.8/63.6/88.9/96% for S1/S2/S3/S4 respectively. Of the 112/134 (84%) of patients whose subtype assignment was stable at the first two timepoints, the mean probability of baseline subtype assignment was 0.93 (Supplementary Fig. 11).

**Figure 4:**
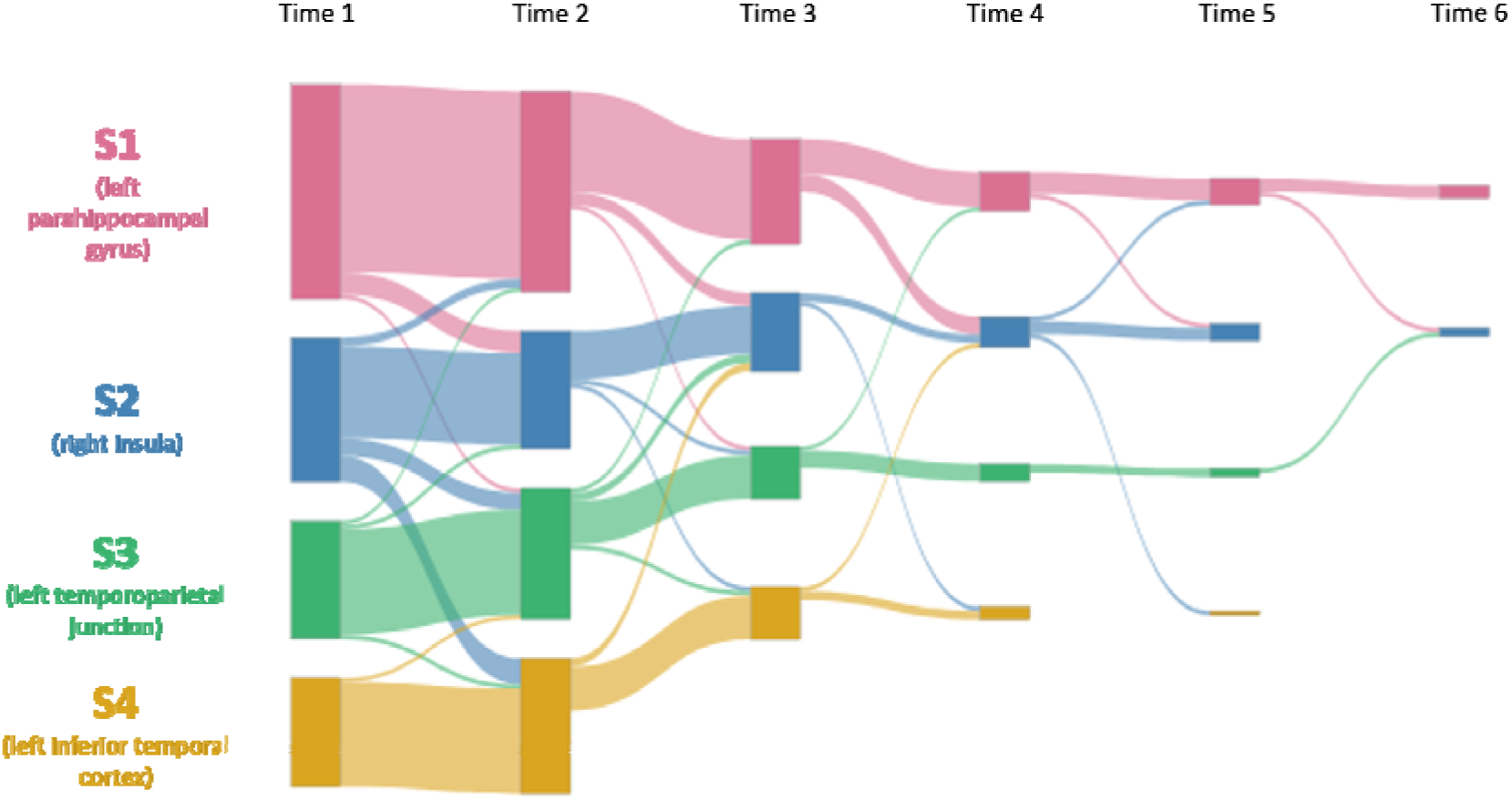
Sankey diagram of subtype assignment between baseline and first follow-up visits. The bars are colour coded to represent the percentage of patients in each subtype at first follow up, stratified by their subtype assignment at baseline S1 left temporal (pink), S2 insula (blue), S3 left TPJ (green), S4 left frontal parietal (yellow).

**Figure 5:**
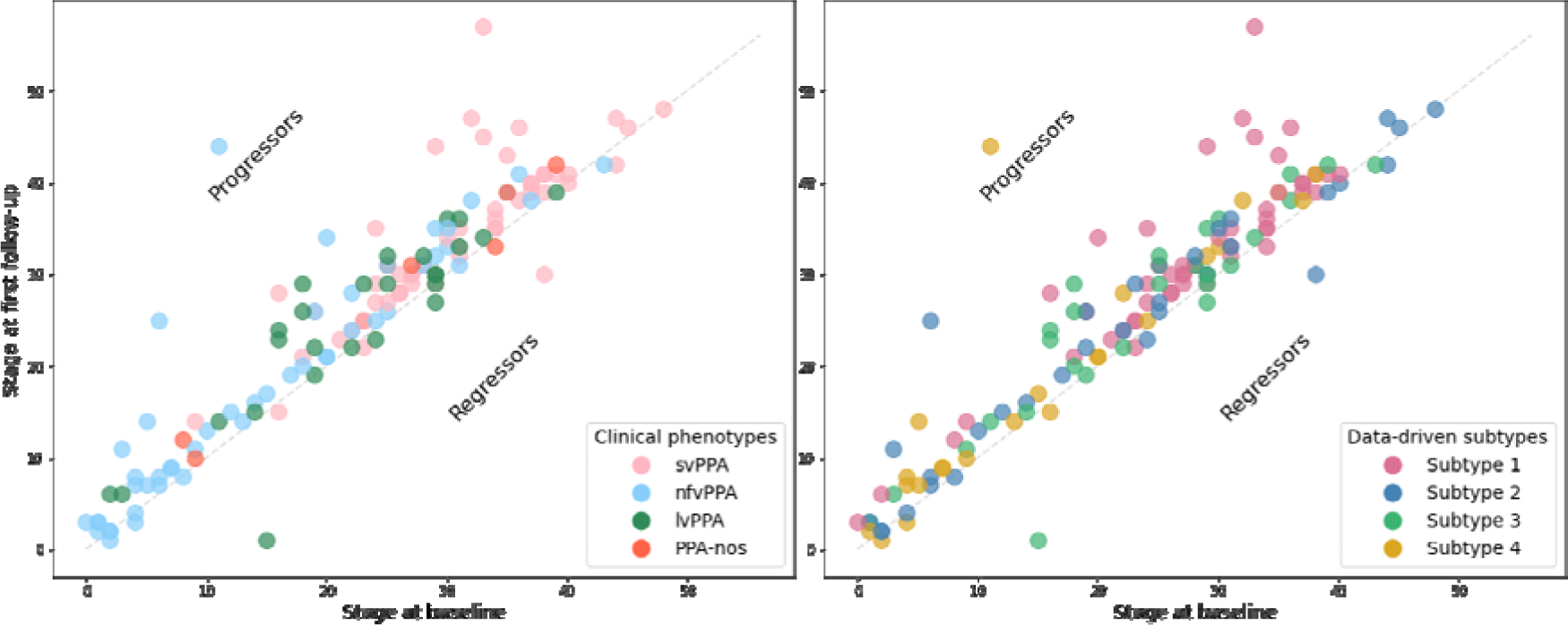
Data-driven stage assignment at baseline and first follow up. In the figure on the left patients are colour coded according to clinical diagnosis, and on the right they are colour coded according to subtype. Those above the diagonal progressed at follow up, whilst those below regressed. Abbreviations: svPPA – semantic variant PPA, nfvPPA – nonfluent/agrammatic variant PPA, lvPPA – logopenic variant PPA, PPA-nos – PPA not otherwise specified.

Figure 4 shows longitudinal consistency of model staging, stratified by model subtype (left) and clinical diagnosis (right). At first follow up, 112/135 (83.0%) patients advanced (upward) to a later stage, 12/135 (8.8%) patients remained at the same stage (on the diagonal), while a further 11/135 (8.1%) patients regressed (downward) to an earlier stage, giving a longitudinal staging consistency exceeding 91.9% (124/135).

Of the 66 patients having three or more MRI scans and subtypable at baseline, 39 were assigned to the same subtype at every timepoint. Amongst these 66 patients, 49 were assigned to monotonically increasing stages across the timepoints.

The analysis of the subset of patients scanned on the same MRI scanner type longitudinally were consistent with the analysis of the full dataset (Supplementary Fig. 13-14).

### Results of ALLFTD test dataset

In the test set five patients were assigned to data driven stage zero, and hence excluded from subsequent analysis. Of those who were subtypable 23 patients were assigned to S1, with an age at first scan of 63.1±6.2 years, 15 patients were assigned to S2, with an age at first scan of 63.2±6.28 years, 9 patients were assigned to S3, with an age at first scan of 68.6±6.4 years, and 14 patients were assigned to S4, with an age at first scan of 66.6±8.1 years.

Supplementary Fig. 15 demonstrates association between clinical variant and subtype assignment in the ALLFTD test set. As in the Queen Square discovery dataset, the greatest phenotype subtype concordance was between svPPA and S1 (left parahippocampal gyrus) (54.1% of svPPA were assigned to S1).

Supplementary Fig.s 16 and 17 demonstrate the longitudinal stability of subtype assignment in the ALLFTD test set. As in the Queen Square discovery dataset, subtypes and stages were stable at follow up clinic visits, with over 90% of individuals remaining in the same stage, or advancing to a later stage at first follow up.

## Discussion

This study aimed to understand neuroanatomical disease progression in PPA, and specifically how neuroanatomical subtypes relate to clinical phenotypes. Our retrospective cohort was one of the largest PPA datasets compiled to date and included individuals with the three named clinical variants as well as PPA-nos. Our model identified four statistically different trajectories of PPA disease progression based on volumetric markers extracted from MRI scans. We also examined disease progression per clinical variant (in a different manner to previous studies ^3,14^), ultimately contributing a unique data-driven perspective on relationships between clinical phenotype and spatiotemporal neuroanatomical pathology subtypes in PPA.

There was a strong correspondence between S1 and svPPA, with the disease progression profile of S1 closely matching the reported patterns of initial temporal pole atrophy in svPPA,^34,42^ evolving to implicate the contralateral hemisphere later in the disease.^43^ In particular, the sequence of decline in S1 overlapped with stages of atrophy reported in TDP-43 Type C, comprising left temporal pole, then the left insula and then the right insula.^13^ Individuals assigned to S1 were largely assigned to stages midway through the disease progression, even at baseline. This reflects the fact that diagnosis typically occurs later for individuals with PPA compared to typical Alzheimer’s disease,^44,45^ particularly in svPPA^10,46^, hence exhibiting relatively advanced accumulation of brain differences at baseline. A side effect is that analyses of atrophy in svPPA struggle to distinguish the very earliest regions of change,^13,47^ as seen in S1 where initial progression through the left temporal pole regions were all staged together. Despite this, S1 was the clearest and most consistent subtype across analyses, matching the notion that svPPA is the most coherent PPA variant clinically, pathologically, and neuroanatomically.^2^

Phenotype-subtype correspondence of the other subtypes was more complicated. The most pronounced correspondence was between S3 and lvPPA – S3 had a high statistical similarity to the disease progression seen in the single phenotype lvPPA model, and 60.8% of patients with a lvPPA diagnosis were assigned to it. Of the data driven subtypes, the sequence of atrophy changes in S3 was more scattered across brain regions, aligning with research that atrophy in lvPPA presents as more diffuse.^34^ However, S2 was also associated with lvPPA (alongside nfvPPA, which had associations with all subtypes). Recent studies exploring heterogeneity in lvPPA have proposed that both the syndrome itself and the sequence of neuroanatomical changes should be understood in a multidimensional manner,^48^ which coincides with the absence of a single lvPPA-specific neuroanatomy amongst the data-driven subtypes in our model. Results from the Queen Square dataset suggest S4 could represent a neuroanatomical subtype of nfvPPA, this subtype was associated with initial left lateralised atrophy of the frontal and temporal lobes regions, which coincides with neuroanatomy reported in a predominantly agrammatic presentation of PPA.^36^

Our discoveries, revealing the presence of numerous neuroanatomical profiles within both lvPPA and nfvPPA, as well as intra-phenotypic overlap, align with the emerging theory that PPA might be more accurately perceived as a spectrum of disorders^36,49^. This perspective is supported by previous PPA research showing that single neuroanatomical profiles can be linked to multiple clinical phenotypes, whilst canonical phenotypes have been observed to align with diverse neuroanatomical patterns,^12^ – as evidenced by up to 62% of individuals with PPA displaying phenotypic attributes that correspond to multiple distinct syndromes.^49^

Regarding the association between neuroanatomical subtypes and PPA-nos, no clear pattern emerged. This may be because the label of PPA-nos encompasses heterogenous disease presentations and potentially diverse neuroanatomical profiles. ^8^ However, it could also stem from the challenge of identifying data-driven patterns within a relatively small sample size (n=16). The number of individuals diagnosed with PPA-nos in the cohort was low considering recent findings indicating that as many as 40% of PPA diagnoses might not align with the clinical consensus criteria for any of the three canonical PPA variants.^8,50^ Prospective research in large, well-characterised cohorts of PPA-nos participants will be required to fully tease apart the neuroanatomical profiles of these cases.

The stability of our PPA subtypes model was consistent over time and across two distinct datasets, underscoring the robustness of the identified subtypes. Furthermore our analysis was robust to excluding pre-2011 data, and also excluding individuals whose scanner type varied longitudinally. ^1,5^

### Limitations and future work

Here we have reported on a retrospective analysis of one of the largest PPA datasets compiled to date. However, we note several limitations that suggest opportunities for future research. Model certainty and results would be improved by having larger sample sizes still, which would facilitate investigation into whether the positional variance of events in the disease progression is reflective of genuine variation in neuroanatomy versus an underpowered sample size. Incorporating neuropsychological and other biomarker data would extend our ability to analyse further subtype distinctions. The absence of participant education levels or ethnicity details makes it challenging to gauge the sample’s representativeness within the broader population of individuals diagnosed with PPA. Moreover, genetic FTD could account for some of the heterogeneity, as previous research has found that neuroanatomic profiles vary between the hereditary and sporadic variants: for example there is greater posterior involvement in genetic nfvPPA^21^. Future work should seek to validate these findings prospectively in large multimodal datasets that are likely to require multi-centre, international collaboration.

The neuroanatomical subtypes may have been influenced by the choice of 19 ROIs included in the model. The choice of brain regions was motivated by wanting to conduct a fine-grained analysis of regions, whilst being constrained by modelling parameters. Further work could leverage clinical knowledge to choose a different subset of ROIs for analysis. Higher resolution datasets may offer opportunities to analyse smaller regions which would have been difficult to reliably parcellate in our dataset.

From the computational perspective, a clustering algorithm might be too blunt of a tool to understand the heterogeneity in pathology underpinning the PPA phenotypes — even a clustering algorithm like SuStaIn that takes into account disease accumulation. Future work should seek to develop models that can explicitly model mixtures of subtypes, and how disease progression diverges and converges across subtypes. We envisage that such a model would be particularly useful for understanding potential similarities between heterogeneous syndromes, which could provide insights into how disease modifying treatments could be used across different phenotypes.

## Conclusion

In this study we identified four distinct patterns of neuroanatomy in clinically diagnosed PPA, with a clear correspondence between S1 and svPPA, and a more variable association between S2/S3/S4 and nfvPPA/lvPPA. This imperfect phenotype–subtype association supports recent conceptualisations of PPA as more appropriately being understood as a spectrum of disease, as opposed to discrete conditions. However, we note that clinical diagnostic labels are undoubtedly helpful and informative for patients and caregivers. Understanding the neuroanatomical, molecular, clinical and cognitive profile of PPA will be crucial for tracking disease progression, screening, and stratification for clinical trials, and developing disease modifying drug treatments. Whilst our work benefitted from one of the largest PPA datasets assembled to date, future work should focus on validating these findings in other datasets with additional data modalities, facilitating further comparison of subtype distinctions.

## Acknowledgements

Data collection and dissemination of the data presented in this manuscript was supported by the ALLFTD Consortium (U19: AG063911, funded by the National Institute on Aging and the National Institute of Neurological Diseases and Stroke) and the former ARTFL & LEFFTDS Consortia (ARTFL: U54 NS092089, funded by the National Institute of Neurological Diseases and Stroke and National Center for Advancing Translational Sciences; LEFFTDS: U01 AG045390, funded by the National Institute on Aging and the National Institute of Neurological Diseases and Stroke). The authors acknowledge the invaluable contributions of the study participants and families as well as the assistance of the support staffs at each of the participating sites.

## Supplementary material

Supplementary material is available in a separate document.

