## Supplementary for "Data-driven neuroanatomical subtypes of primary progressive aphasia"

Supplementary material

### Queen Square discovery dataset

#### Baseline scans

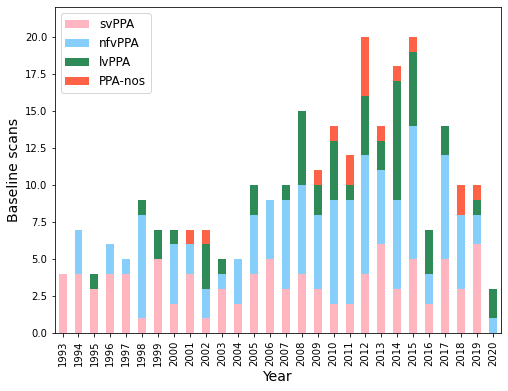

Supplementary Figure 1: **Bar chart of baseline scans by year.**

Abbreviations: svPPA – semantic variant PPA, nfvPPA – nonfluent/agrammatic variant PPA, lvPPA – logopenic variant PPA, PPA-nos – PPA not otherwise specified.

Baseline and follow-up data was collected from participants in five FTD studies at the UCL Queen Square Institute of Neurology Dementia Research Centre between 1993 and 2020. These were: Brain signatures of auditory information processing in the degenerative dementias (06N032, approved by The National Hospital for Neurology and Neurosurgery & UCL Institute of Neurology Joint Research Ethics Committee); Familial and sporadic non-Alzheimer degenerative dementia: longitudinal clinical, biochemical and neuroimaging studies (02/N099, approved by The National Hospital for Neurology and Neurosurgery & UCL Institute of Neurology Joint Research Ethics Committee); Longitudinal Investigation of Fronto-Temporal Dementia and associated disorders (LIFTD) (150805, approved by University College London Hospitals & University College London Joint Research Office); The burden of frontotemporal dementia: clinical course, outcome and caregiving (97/N077, approved by Joint Medical Ethics Committee, The National Hospital for Neurology & Neurosurgery); Impaired language output in frontotemporal lobar degeneration: a cross-sectional and longitudinal clinical, neuropsychological and neuroimaging study (05/Q0512/99, approved by The National Hospital for Neurology and Neurosurgery & UCL Institute of Neurology Joint Research Ethics Committee).

#### Scanner types

Supplementary Table 1 details the three different types of MRI scanners, and acquisition parameters used to acquire the images in the Queen Square discovery dataset.

| Type | 3T Trio | 3T Prisma | 1.5T Signa |
| --- | --- | --- | --- |
| Vendor | Siemens, Erlangen, Germany | Siemens, Erlangen, Germany | GE Medical systems, Milwaukee, WI |
| Scans in study (patients:controls) | 114:46 | 42:27 | 114:48 |
| Repetition time (ms) | 2200 | 2000 | 12 |
| Inversion time (ms) | 900 | 850 | 650 |
| Echo time (ms) | 2.9 | 2.93 | 5 |
| Acquisition matrix | 256x256 | 256x256 | 256x256 |
| Spatial resolution (mm) | 1.1 | 1.1 | 1.5 |

Supplementary Table 1: Scanners used in data collection of Queen Square discovery dataset.

#### Scanner types ALLFTD

Supplementary Table 2 details the three different types of MRI scanners, and acquisition parameters used to acquire the images in the ALLFTD test dataset.

| Type | 3T Trio | 3T Signa |
| --- | --- | --- |
| Vendor | Siemens, Erlangen, Germany | GE Medical systems, Milwaukee, WI |
| Scans in study (patients:controls) | 65:111 | 1:6 |
| Repetition time (s) | 2.3 | 2.3 |
| Inversion time (s) | 0.9 | 0.9 |
| Echo time (ms) | 2.98 | 2.98 |
| Acquisition matrix | 160x160 | 166x166 |
| Spatial resolution (mm) | 1.2 | 1.2 |

Supplementary Table 2: Scanners used in data collection of Queen Square discovery dataset.

### Regions of interest

Supplementary Table 3 records the 19 brain regions of interest (ROIs) included in the model.

|  |  | ROI | Sub-regions | Queen Square discovery dataset | ALLFTD test dataset |
| --- | --- | --- | --- | --- | --- |
|  |  |  |  | w-score | w-score |
| 1 | Temporal Lobe | Left temporal pole |  | -2.5 (2.0) | -2.5 (2.2) |
| 2 |  | Left superior temporal gyrus |  | -2.1 (1.4) | -1.2 (1.3) |
| 3 |  | Left middle temporal gyrus |  | -2.8 (1.8) | -1.9 (1.8) |
| 4 |  | Left inferior temporal cortex | Inferior temporal gyrus | -3.5 (2.6) | -2.6 (2.4) |
|  |  |  | Fusiform gyrus |  |  |
| 5 |  | Left amygdala |  | -3.0 (2.3) | -2.9 (2.3) |
| 6 |  | Left parahippocampal gyrus | Parahippocampal gyrus | -1.9 (1.7) | -3.0 (2.6) |
|  |  |  | Entorhinal areas |  |  |
| 7 |  | Left hippocampus |  | -1.9 (1.7) | -1.6 (1.4) |
| 8 |  | Right temporal pole |  | -1.2 (1.6) | -1.8 (2.0) |
| 9 |  | Right middle temporal gyrus |  | -1.5 (1.7) | -1.3 (1.7) |
| 10 |  | Right inferior temporal cortex | Inferior temporal gyrus | -1.5 (1.8) | -1.8 (2.4) |
|  |  |  | Fusiform gyrus |  |  |
| 11 |  | Right amygdala |  | -1.6 (1.7) | -1.7 (1.9) |
| 12 | Parietal Lobe | Left temporoparietal junction | Supramarginal gyrus | -1.7 (1.6) | -0.7 (0.8) |
|  |  |  | Angular gyrus |  |  |
|  |  |  | Planum temporale |  |  |
| 13 |  | Left other parietal | Parietal operculum | -1.2 (1.5) | --0.0 (1.0) |
|  |  |  | Superior parietal lobule |  |  |
|  |  |  | Postcentral gyrus |  |  |
|  |  |  | Postcentral gyrus medial segment |  |  |
| 14 | Frontal Lobe | Left inferior frontal gyrus |  | -1.2 (1.5) | -2.5 (2.3) |
| 15 |  | Left other frontal | Superior frontal gyrus | -1.3 (1.7) | -0.6 (1.6) |
|  |  |  | Middle frontal gyrus |  |  |
|  |  |  | Medial frontal cortex |  |  |
|  |  |  | Subcallosal area |  |  |
|  |  |  | Superior frontal gyrus medial segment |  |  |
|  |  |  | Precentral gyrus medial segment |  |  |
|  |  |  | Frontal operculum |  |  |
|  |  |  | Central operculum |  |  |
|  |  |  | Frontal pole |  |  |
| 16 |  | Left precentral gyrus |  | -0.9 (1.3) | -0.4 (1.4) |
| 17 |  | Left anterior cingulate |  | -1.4 (1.7) | -0.8 (1.4) |
| 18 | Insula cortex | Left insula | Posterior insula | -2.9 (1.8) | -2.8 (1.7) |
|  |  |  | Anterior insula |  |  |
| 19 |  | Right insula | Posterior insula | -1.7 (1.5) | -1.5 (1.8) |
|  |  |  | Anterior insula |  |  |

Supplementary Table 3: **Regions of interest included in the model.** The 19 regions included in the model, as well as a breakdown of sub regions within them. The w-scores result from standardising the raw volumes (calculated using geodesic information flow (GIF)^1^ and the neuromorphometrics brain parcellation) with respect to covariates of: age, sex, total intercranial volume and scanner type. The w-scores for the Queen Square dataset and ALLFTD dataset are calculated with respect to controls from the corresponding dataset.

### Model hyperparameters

#### w-score thresholds

Supplementary Figure 2 shows the cumulative distribution function of w-scores for each of the 19 regions of interest in the Queen Square dataset. The model used three severity scores (1,2,3) as thresholds of abnormality from the control population.^2^ We justified the choice of three severity scores since the 95^th^ percentile falls beyond a w-score of three in all 19 regions, so at least 5% of the sample have reached the most severe score in all regions.^3^

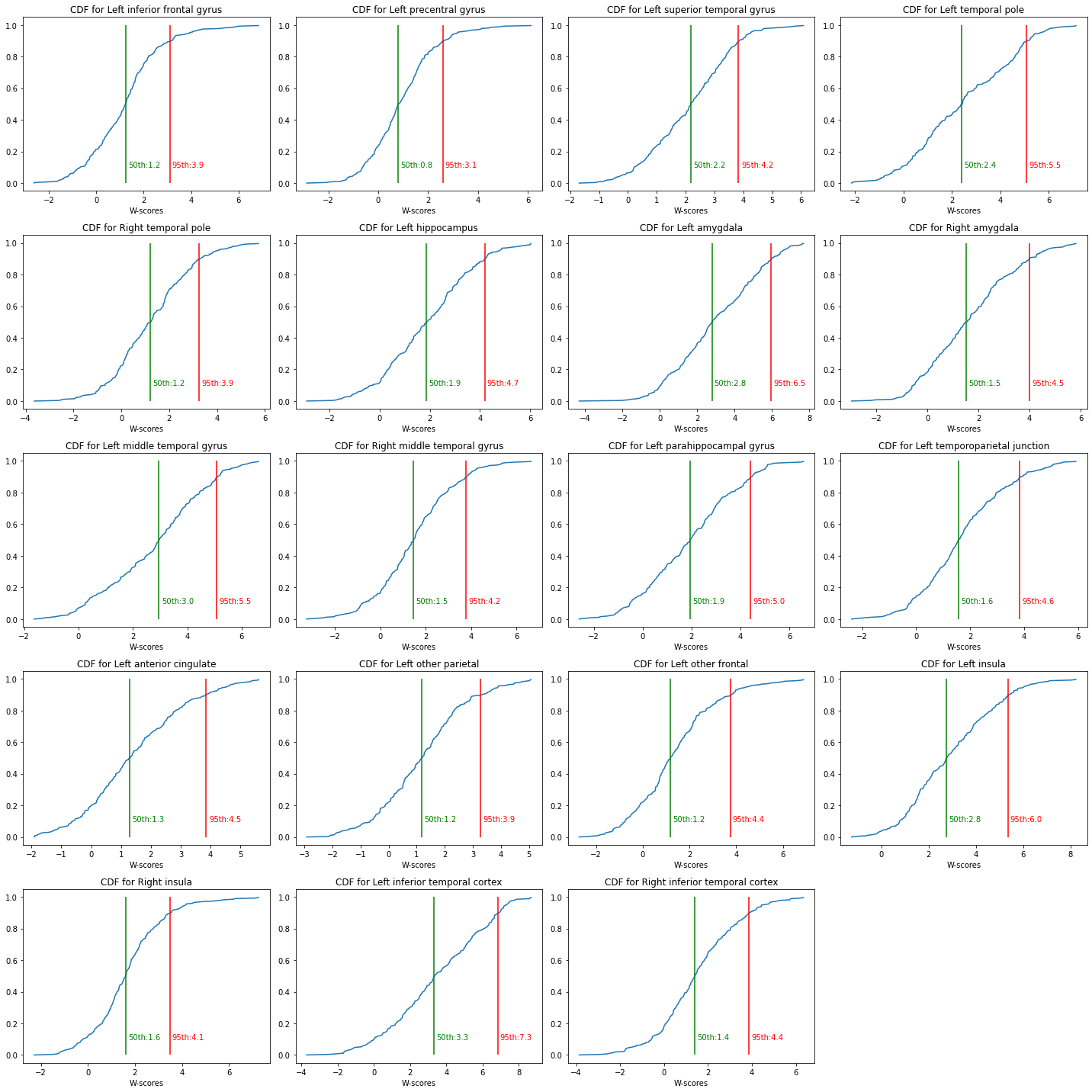

Supplementary Figure 2: **Cumulative distribution function of w-scores across the 19 ROIs** used in the model. Green vertical line represents the 50^th^ percentile of each region (50th: w-score corresponding to 50th percentile), red vertical line represents the 95th percentile of each region (95th: w-score corresponding to 95th percentile).

#### Model fitting

Supplementary Figure 3a demonstrates the test set log likelihood across folds, Supplementary Figure 3b demonstrates the cross-validation information criterion (CVIC) achieved by the model for n subtypes displayed along the x-axis.^2^ Higher log-likelihood and lower CVIC represents better model fit.

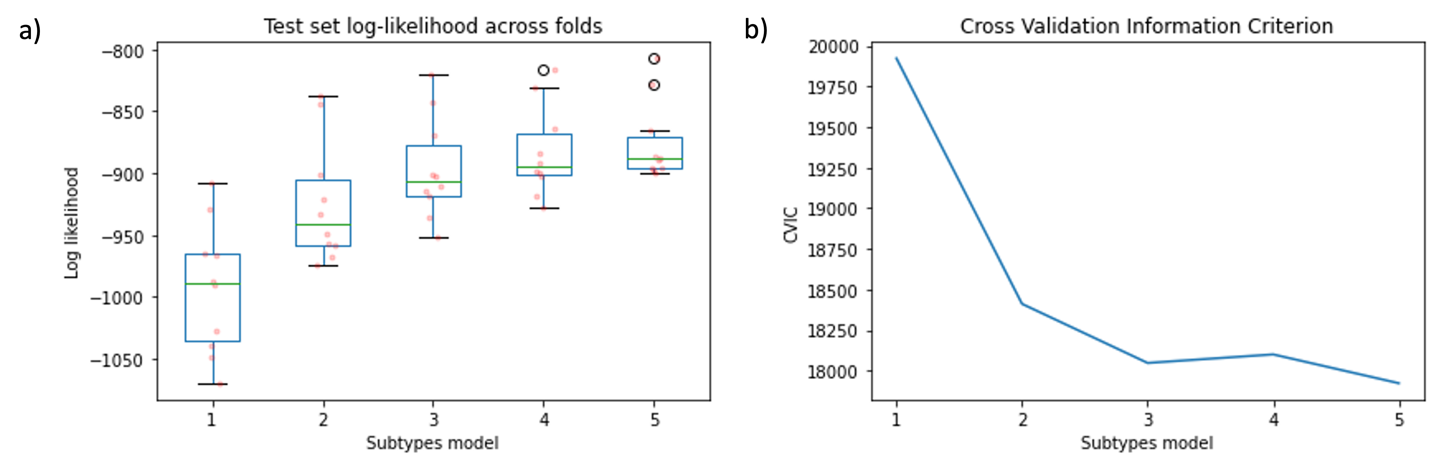

Supplementary Figure 3: a) Test set log likelihood across folds b) Cross validation information criterion for n subtype models.

#### Hellinger distance similarity measure for data-driven subtypes

To have a metric of statistical similarity between positional variance diagrams we calculated a Hellinger distance reference value. Following the framework outlined by Oxtoby *et al*^4^ for each of the four model subtypes, we calculated the mean Hellinger distance between a new positional variance diagram (resulting from randomly permuting its rows), and the other three subtypes. For each subtype we repeated this Hellinger distance of permuted positional variance 100 times, the mean across all values was taken to be the reference value $H_{0}=0.89\pm0.019$ (95% CI [0.893, 0.895]).

### Results: Queen Square discovery set

#### Stage and subtype assignment

Supplementary Figure 4 shows the number of individuals assigned each stage by subtype. The model included 57 stages, but no patients were above stage 50, with most patients assigned to early and mid-stages of the disease.

Patients were assigned a subtype according to the choice which maximised their subtype probability. Following Vogel *at al*^5^ Supplementary Figure 5 visualises the distribution of these probabilities using a quaternary plot. Each point (representing one baseline data point) is located within the pyramid using the probability of subtype assignment to determine its distance from the corresponding subtype vertex. As such, those points representing patients with high certainty of subtype assignment are located near to subtype vertices, whilst patients with more variable subtype assignment are located near the middle of the pyramid. This same data is shown in a boxplot in Supplementary Figure 11.

Supplementary Table 4 shows the paired Hellinger distance between subtypes positional variance diagrams. The low similarity between subtypes supports them representing statistically different sequences of neuroanatomy events.

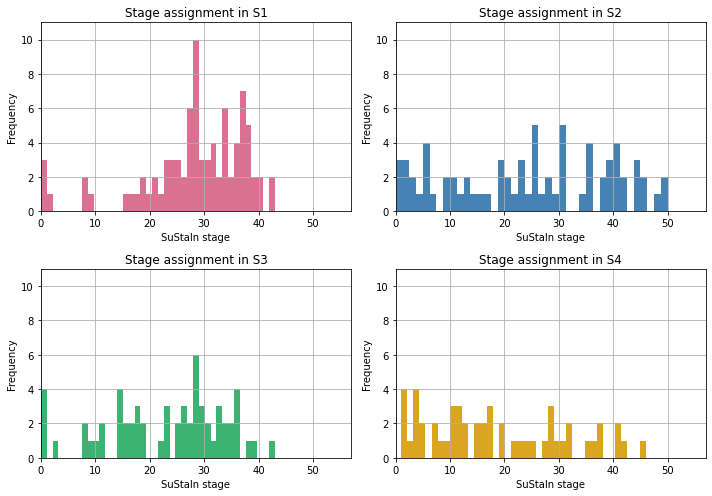

Supplementary Figure 4: **Data-driven stage assignment in the four data-driven subtypes.** Colours are used to distinguish the four subtypes: S1 (pink), S2 (blue), S3 (green), S4 (yellow).

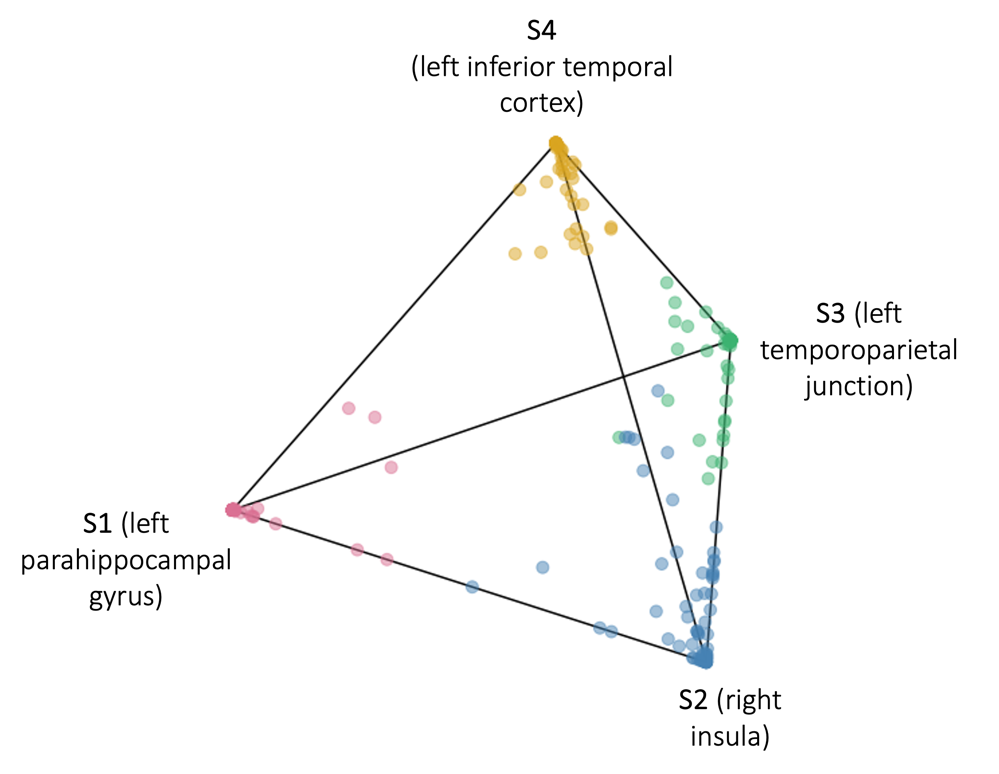

Supplementary Figure 5: **Quaternary plot showing the probability each patient is assigned to each subtype.** Colours are used to distinguish the four subtypes: S1 (pink), S2 (blue), S3 (green), S4 (yellow).

|  | S1  (left parahippocampal gyrus) | S2  (right insula) | S3  (left temporoparietal junction) | S4 (left inferior temporal cortex) |
| --- | --- | --- | --- | --- |
| S1  (left parahippocampal gyrus) | 0.0 | 0.85 | 0.8 | 0.78 |
| S2  (right insula) | 0.85 | 0.0 | 0.82 | 0.9 |
| S3  (left temporoparietal junction) | 0.8 | 0.82 | 0.0 | 0.81 |
| S4  (left inferior temporal cortex) | 0.78 | 0.9 | 0.81 | 0.0 |

Supplementary Table 4: **Paired Hellinger distances between data-driven subtypes**. A Hellinger distance of 0 indicates perfect agreement between two subtypes posterior event distributions, and 1 indicates complete disagreement between two subtypes posterior event distributions. Colours are used to distinguish the four subtypes: S1 (pink), S2 (blue), S3 (green), S4 (yellow).

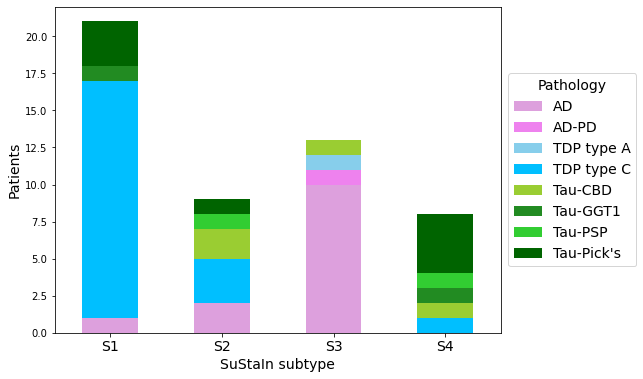

Supplementary Figure 6: **Comparison between data-driven subtype and primary pathology.** Stacked bar chart represents the number of patients with each primary pathology by data-driven subtype.

Abbreviations: AD – Alzheimer’s disease, AD-PD – Alzheimer’s disease with Parkinsons, TDP type A, TDP – TAR DNA binding protein 43, Tau – FTLD-tau.

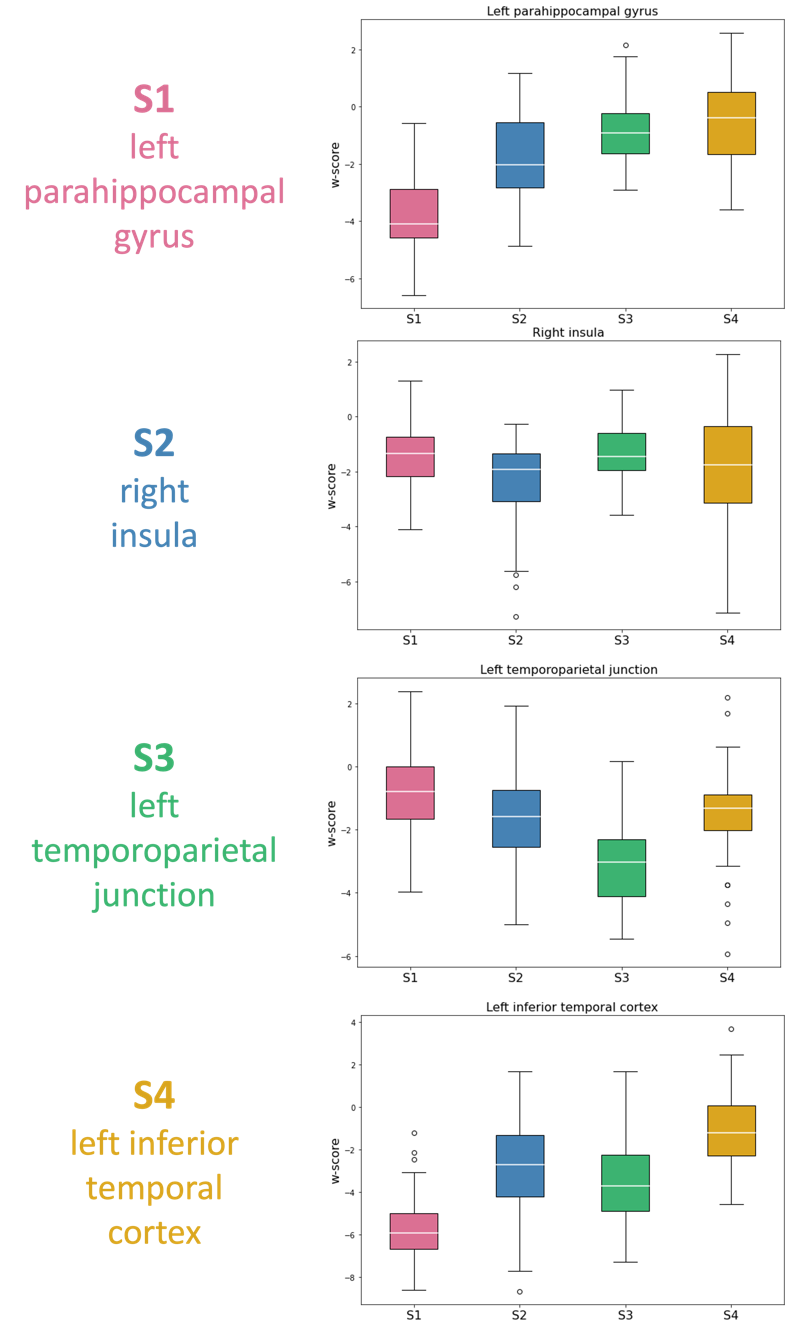

Supplementary Figure 7: **Key regions of brain volume change for each subtype*.*** Each boxplot depicts the area with the most statistically different change in brain volume compared to controls at baseline, for the subtype to the left. For S1/S2/S3/S4 this was the parahippocampal gyrus/ right insula/ temporoparietal junction/ inferior temporal cortex. Colours correspond to subtype assignment at baseline S1 left parahippocampal gyrus (pink), S2 right insula (blue), S3 left temporoparietal junction (green), S4 left inferior temporal cortex (yellow).

#### Association with clinical phenotypes

##### Phenotypic disease progression

Supplementary Figure 8 shows a single disease progression pattern per clinical phenotype. Each disease progression pattern is estimated using SuStaIn (with the subtype parameter set to one – i.e. a fixed single ‘subtype’) with data from a single phenotype.^2,6^

Supplementary Figure 9 shows the single disease progression pattern per clinical phenotype, where the model complexity has been reduced, down to a single normal/abnormal w-score event. Each disease progression pattern is estimated using SuStaIn (with a single ‘subtype’) with data from a single phenotype, but this time using just one w-score of 1. The sequence of ROIs becoming abnormal is very similar to the three w-score model (Supplementary Figure 8). The one-w-score model offers a clearer visualisation of disease progression, which we subsequently utilise for analysis in Supplementary Figure 10.

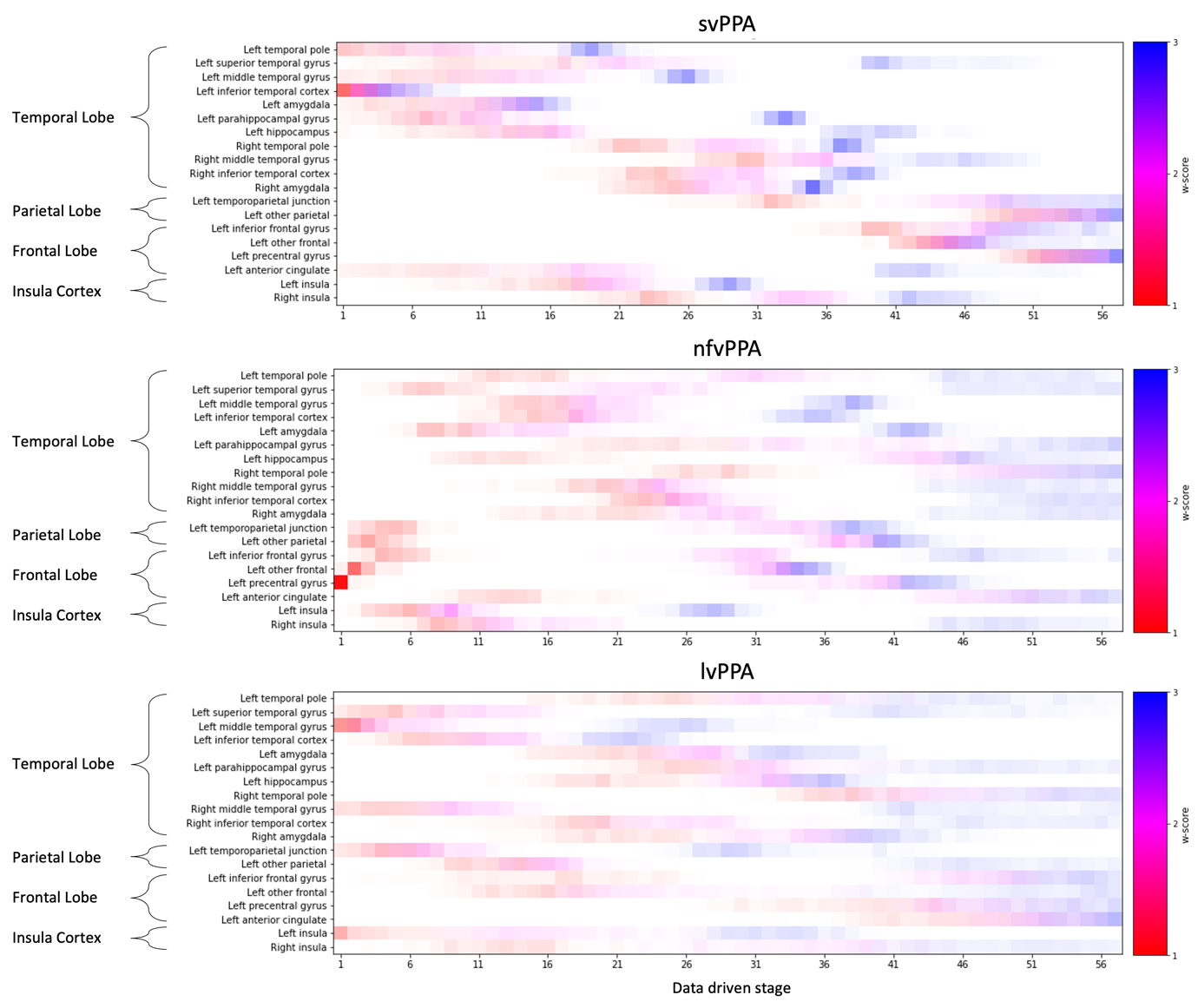

Supplementary Figure 8: **Positional variance diagrams for phenotypic data.** Along the y-axis are the regions of interest used in the model, grouped by location in the brain. The data-driven stages correspond to the sequence that brain regions become abnormal, with colour representing degree of abnormality (w-score 1: red, w-score 2: pink, w-score 3: blue), and colour density representing model certainty.

Abbreviations: svPPA – semantic variant PPA, nfvPPA – non-fluent/agrammatic variant PPA, lvPPA – logopenic variant PPA.

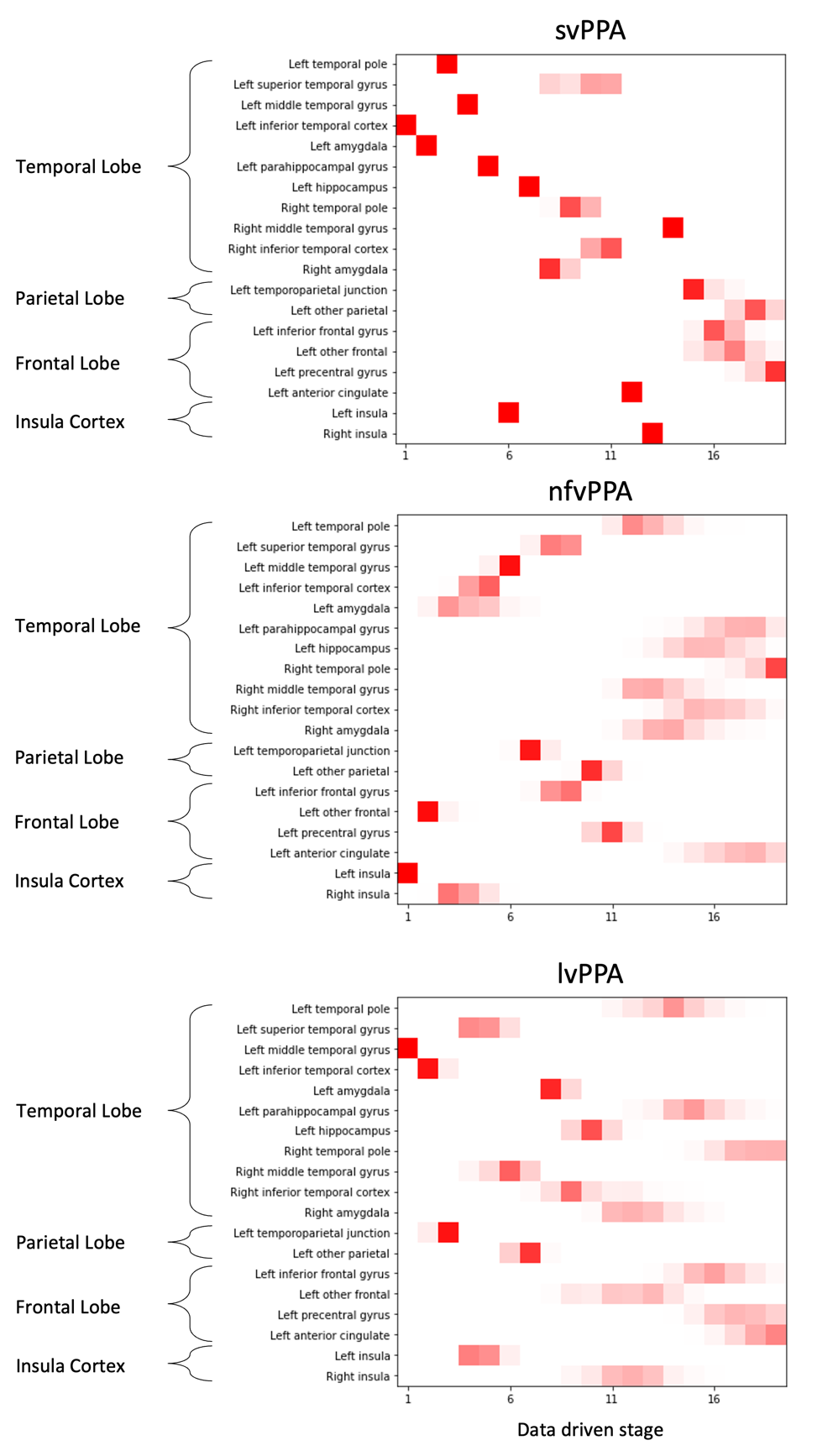

Supplementary Figure 9: **Positional variance diagrams for phenotypic data with one w-score.** Along the y-axis are the regions of interest used in the model, grouped by location in the brain. The data-driven stages correspond to the sequence that brain regions become abnormal, with colour density representing model certainty.

Abbreviations: SuStaIn – Subtype and Stage Inference algorithm, svPPA – semantic variant PPA, nfvPPA – non-fluent/agrammatic variant PPA, lvPPA – logopenic variant PPA.

##### Phenotypic disease progression vs. Subtype disease progression

Supplementary Table 5 records the paired Hellinger distances between the four data-driven subtypes and the three clinical phenotypes.

Supplementary Figure 10 shows the sequence that ROIs become abnormal in each subtype compared to the clinical phenotypes. The sequences of ROI progression in the data-driven subtypes were extracted from the three w-score positional variance diagrams by calculating at which stage each event reached w-score=1 with a probability of 0.75. These stages were then used to order the events sequentially. The sequences were calculated similarly in the clinical phenotypes, but directly from the one w-score model. In the subplots a/b/c/d the x-axis is ordered according to the sequence of disease progression in S1/S2/S3/S4 respectively. Each clinical phenotype is then plotted as a connected line denoting when each event occurred. Complete agreement between sequence of ROIs in the subtype and clinical phenotype would follow the identity line. In the figure we highlight the sequences which were statistically correlated according to the Hellinger distance analysis (Supplementary Table 5): S1 and svPPA (shown in bold pale pink in Supplementary Figure 10a); S3 and lvPPA (shown in bold dark green in Supplementary Figure 10c).

|  | | Data-driven subtype | | | |
| --- | --- | --- | --- | --- | --- |
|  |  | S1  (left parahippocampal gyrus) | S2  (right insula) | S3  (left temporoparietal junction) | S4  (left inferior temporal cortex) |
| Clinical phenotype | svPPA | 0.44 | 0.77 | 0.85 | 0.90 |
|  | nfvPPA | 0.86 | 0.75 | 0.74 | 0.60 |
|  | lvPPA | 0.82 | 0.73 | 0.46 | 0.75 |

Supplementary Table 5: **Hellinger distances between phenotype models and data-driven subtypes.** The Hellinger distance between the posterior identified for each clinical diagnosis compared to the posterior identified for the four data-driven subtypes. Colours are used to distinguish the four subtypes: S1 (pink), S2 (blue), S3 (green), S4 (yellow).

Abbreviations: svPPA – semantic variant PPA, nfvPPA – nonfluent/agrammatic variant PPA, lvPPA – logopenic variant PPA.

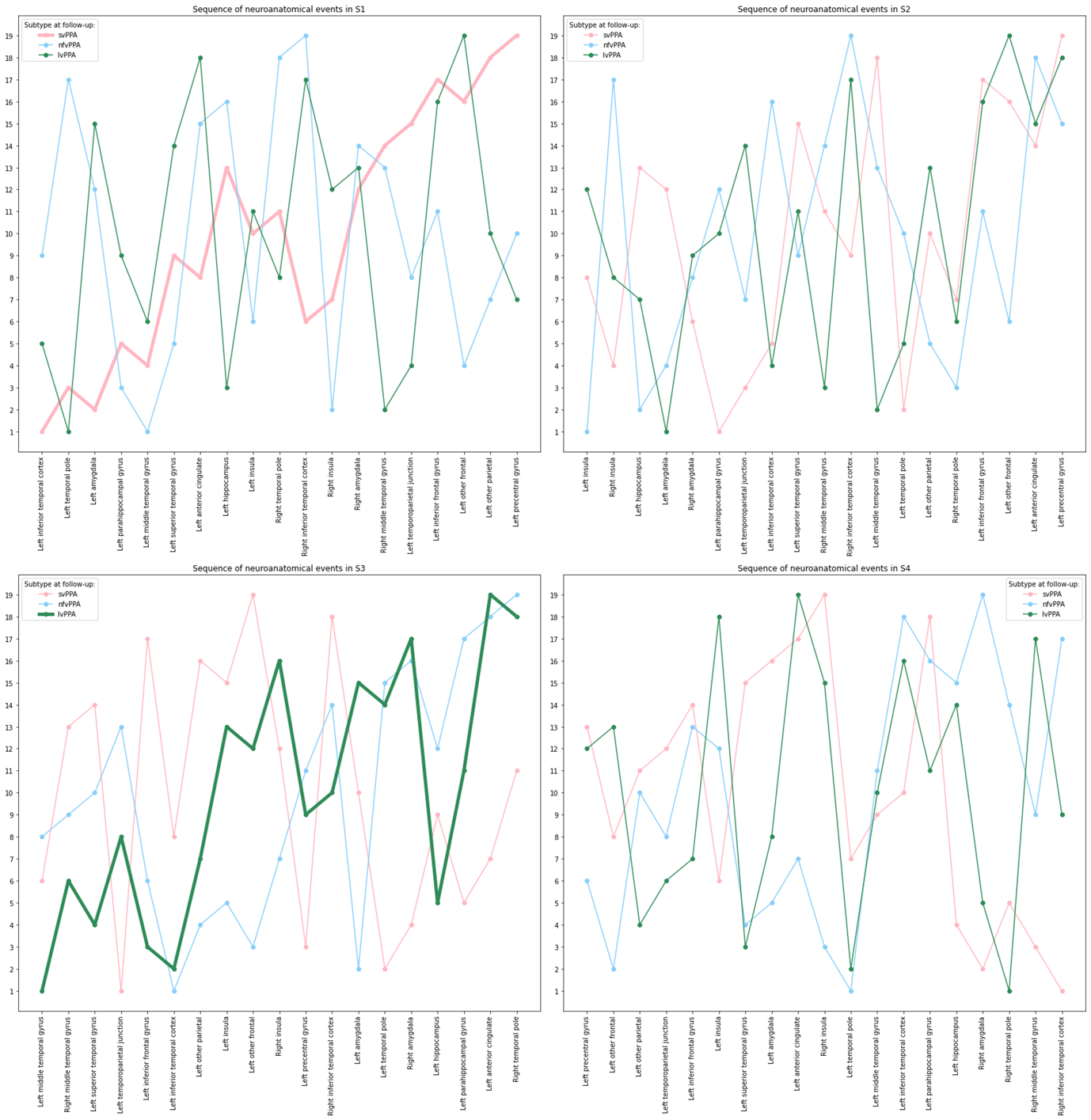

Supplementary Figure 10: **Sequence of neuroanatomical pathology:** **data-driven subtypes vs. phenotypes.** Along the x-axis are the regions of interest used in the model, ordered according to the sequence of neuroanatomical pathology per subtype, a) S1 left parahippocampal gyrus, b) S2 rught insula, c) S3 left temporoparietal junction, d) S4 left inferior temporal cortex. The y-axis is the sequence of neuroanatomical pathology in the clinical phenotypes. Colours are used to distinguish the three phenotypes: svPPA (pale pink), nfvPPA (pale blue), lvPPA (dark green).

Abbreviations: svPPA – semantic variant PPA, nfvPPA – non-fluent/agrammatic variant PPA, lvPPA – logopenic variant PPA.

#### Longitudinal analysis

Supplementary Figure 11a shows the distribution of subtype assignment probabilities at baseline stratified by whether patients changed subtype assignment at first follow up. Supplementary Figure 11b shows the distribution of subtype assignment probabilities at baseline stratified by whether patients regressed or progressed/were stable at first follow up.

Supplementary Figure 12 shows the probability of baseline subtype assignment.

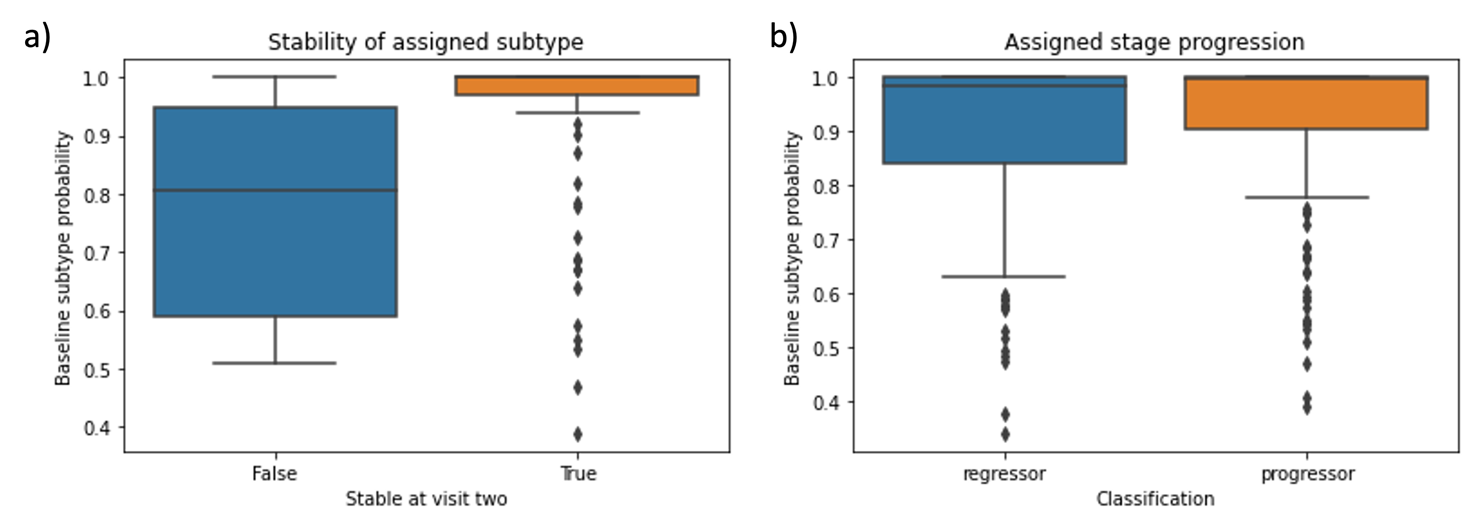

Supplementary Figure 11: a) **Subtype consistency at first follow up visit**. The y-axis shows the baseline subtype probability for the subtype individuals were assigned to, so a value of one suggests high confidence in the subtype assignment, whilst a value nearer to 0 suggests lower confidence. The x-axis labels correspond to whether patients assigned subtype changed at first follow up (False), or stayed the same (True). b) **Staging consistency at first follow up**. The y-axis shows the baseline subtype probability for the subtype individuals were assigned to, so a value of one suggests high confidence in the subtype assignment, whilst a value nearer to 0 suggests lower confidence. The x-axis labels correspond to whether patients assigned stage was consistent at first follow up (progressor – they progressed or remained stable) or inconsistent (they regressed).

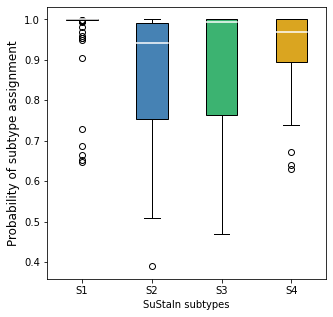

Supplementary Figure 12: **Probability of subtype assignment at baseline across the four subtypes.** The y axis is the baseline probability amongst patients assigned to the respective subtypes at baseline.

#### Longitudinal analysis of stable scanner type

Since 22% (30/137) of individuals changed scanner manufacture or scanner strength longitudinally we repeated the longitudinal analysis restricted to those who were scanned on the same scanner at all time points.

Of the 107 individuals with longitudinal data collected on the same scanner, 105 were deemed subtypable as they were assigned above stage 0 at baseline. Supplementary Figure 13 shows longitudinal consistency of model subtype assignments for these 105 patients. Subtype consistency between first and second MRI scan was: 89.5/60.7/95.2/94.4% for S1/S2/S3/S4 respectively. Of the 88/105 (83.8%) of patients whose subtype assignment was stable at the first two timepoints, the mean probability of baseline subtype assignment was 0.93.

Supplementary figure 14 shows longitudinal consistency of model staging, stratified by model subtype (left) and clinical diagnosis (right). At first follow up, 87/105 (82.9%) patients advanced (upward) to a later stage, 12/105 (11.4%) patients remained at the same stage (on the diagonal), while a further 6/105 (5.7%) patients regressed (downward) to an earlier stage, giving a longitudinal staging consistency exceeding 94.3% (99/105).

Of the 42 patients having three or more MRI scans and subtypable at baseline, 28 were assigned the same subtype at every timepoint, and 33 were assigned monotonically increasing stages across the timepoints.

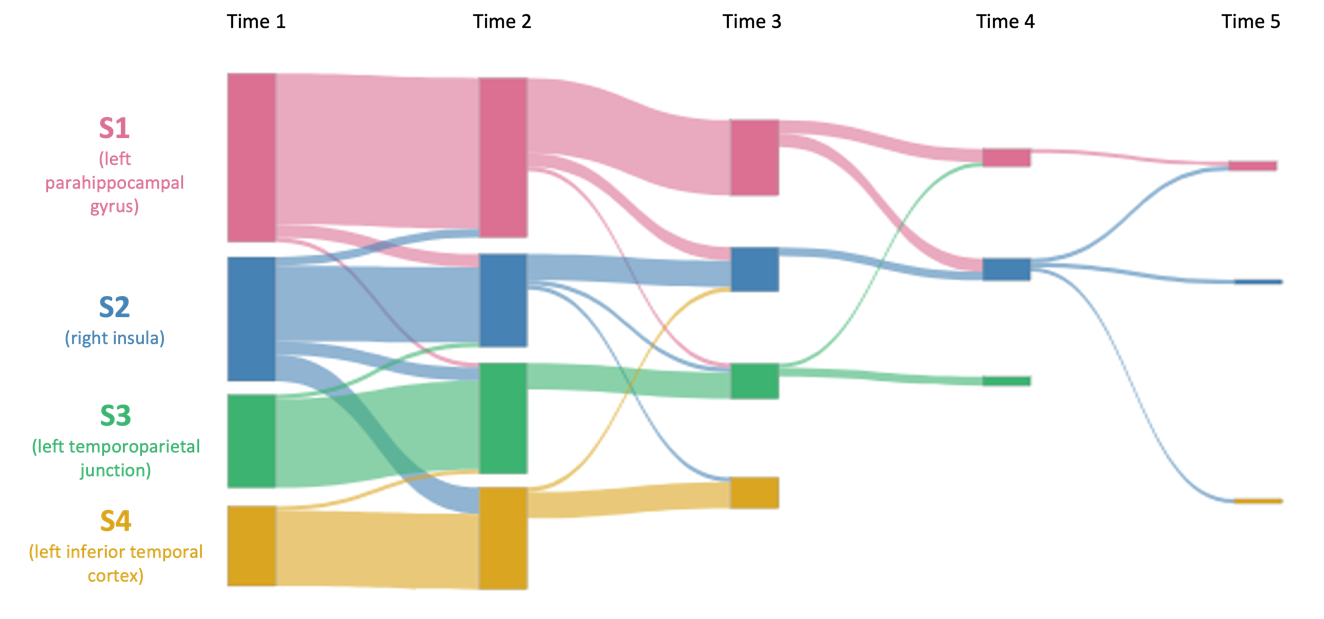

Supplementary Figure 13: **Sankey diagram of subtype assignment between baseline and first follow-up clinic visit.** The bars are colour coded to represent the percentage of patients in each subtype at first follow up, stratified by their subtype assignment at baseline S1 anterior temporal (pink), S2 insula (blue), S3 TPJ (green), S4 precentral gyrus (yellow). The data was restricted to just those individuals at each time who had been scanned on the same type of scanner up to and including that time.

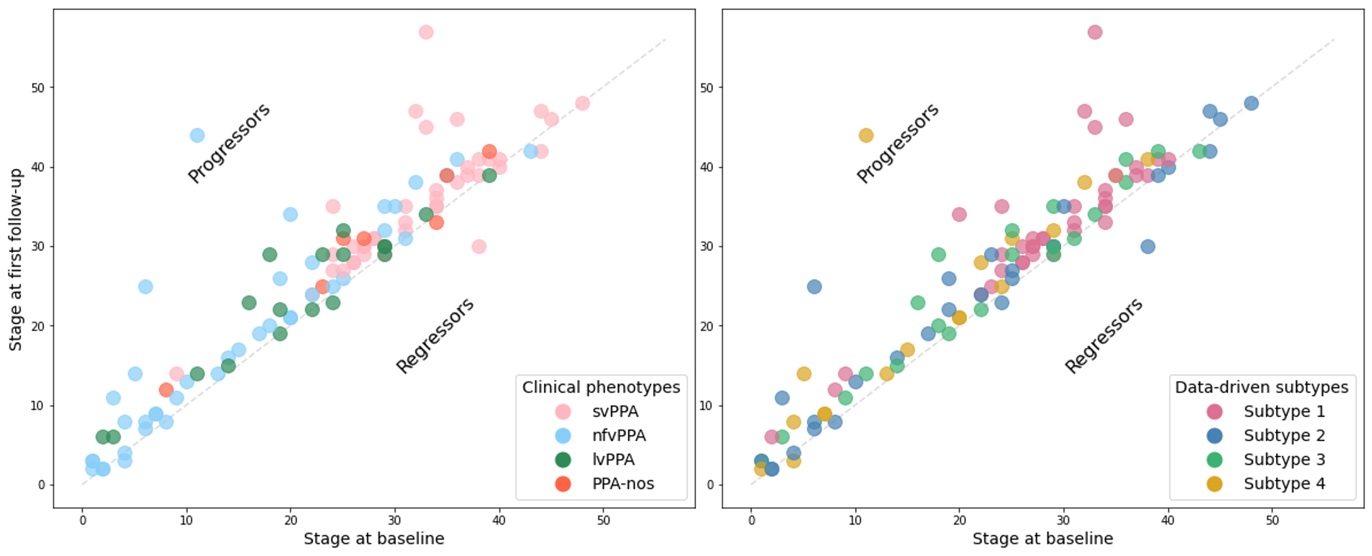

Supplementary Figure 14: **Data-driven stage assignment at baseline and first follow up.** In the figure on the left patients are colour coded according to clinical diagnosis, and on the right they are colour coded according to subtype. Those above the diagonal progressed at follow up, whilst those below regressed.

Abbreviations: svPPA – semantic variant PPA, nfvPPA – nonfluent/agrammatic variant PPA, lvPPA – logopenic variant PPA, PPA-nos – PPA not otherwise specified.

### Results: ALLFTD test set

#### ALLFTD association with clinical phenotypes

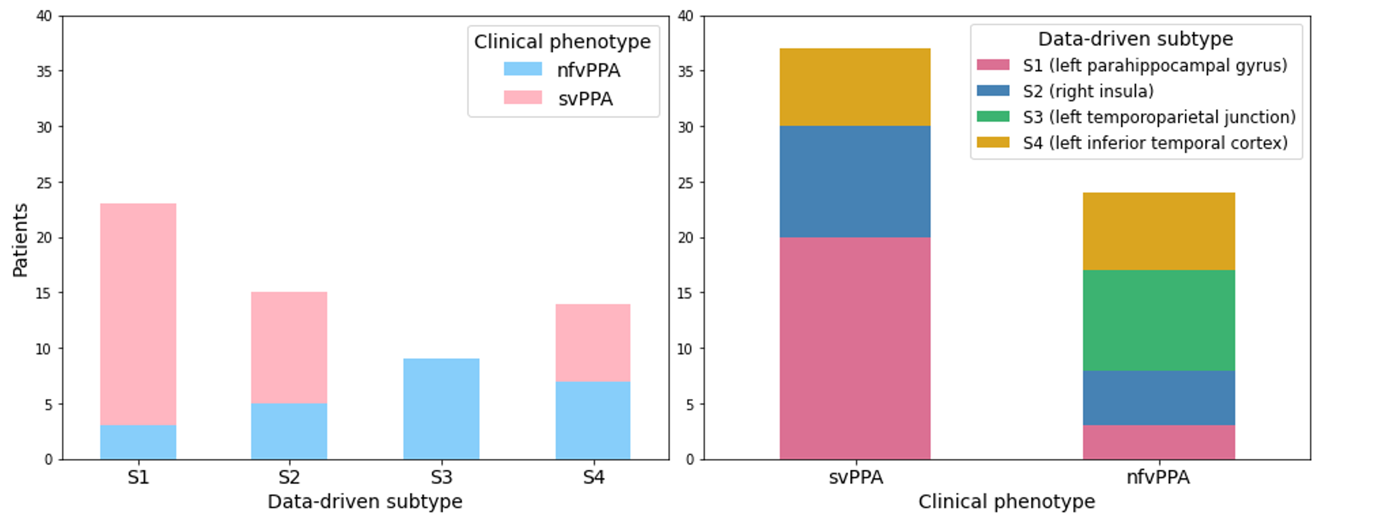

Supplementary Figure 15: **Comparison between data-driven subtype assignment and clinical diagnosis in ALLFTD dataset.** The stacked bar chart on the left shows the number of patients with each clinical diagnosis by data-driven subtype assignment. The figure on the right shows the number of patients who were assigned each data-driven subtype by clinical diagnosis.

Abbreviations: svPPA – semantic variant PPA, nfvPPA – nonfluent/agrammatic variant PPA.

#### ALLFTD Longitudinal analysis

Supplementary Figure 16 is a Sankey diagram demonstrating the longitudinal assignment to subtype for the 58 patients who had longitudinal data and were subtypable at baseline.

Supplementary Figure 17 shows longitudinal consistency of model staging, stratified by model subtype (left) and clinical diagnosis (right). At first follow up, 46/53 (86.8%) patients advanced (upward) to a later stage, 4/53 (7.5%) patients remained at the same stage (on the diagonal), while a further 3/53 (5.7%) patients regressed (downward) to an earlier stage, giving a longitudinal staging consistency exceeding 94.3% (50/53).

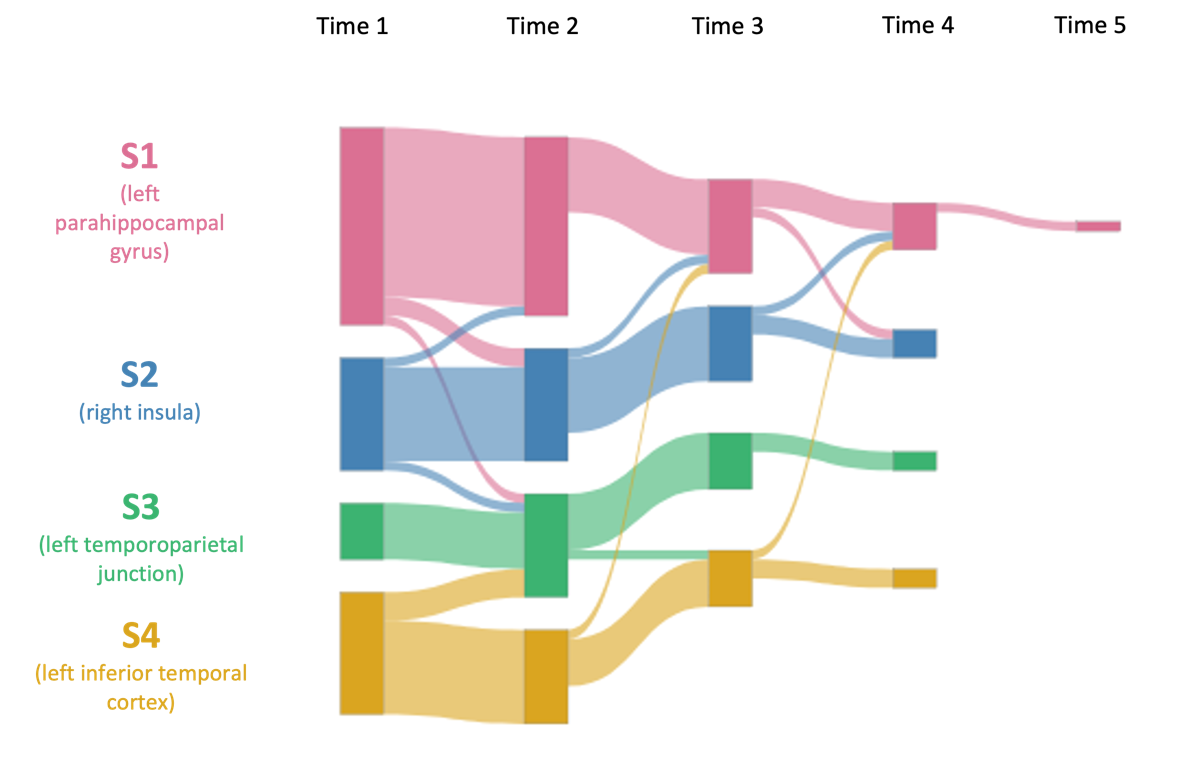

Supplementary Figure 16: **Sankey diagram of subtype assignment between baseline and follow-up visits in ALLFTD test set.** The bars are colour coded to represent the percentage of patients in each subtype at first follow up, stratified by their subtype assignment at baseline S1 left temporal (pink), S2 insula (blue), S3 left TPJ (green), S4 left frontal parietal (yellow).

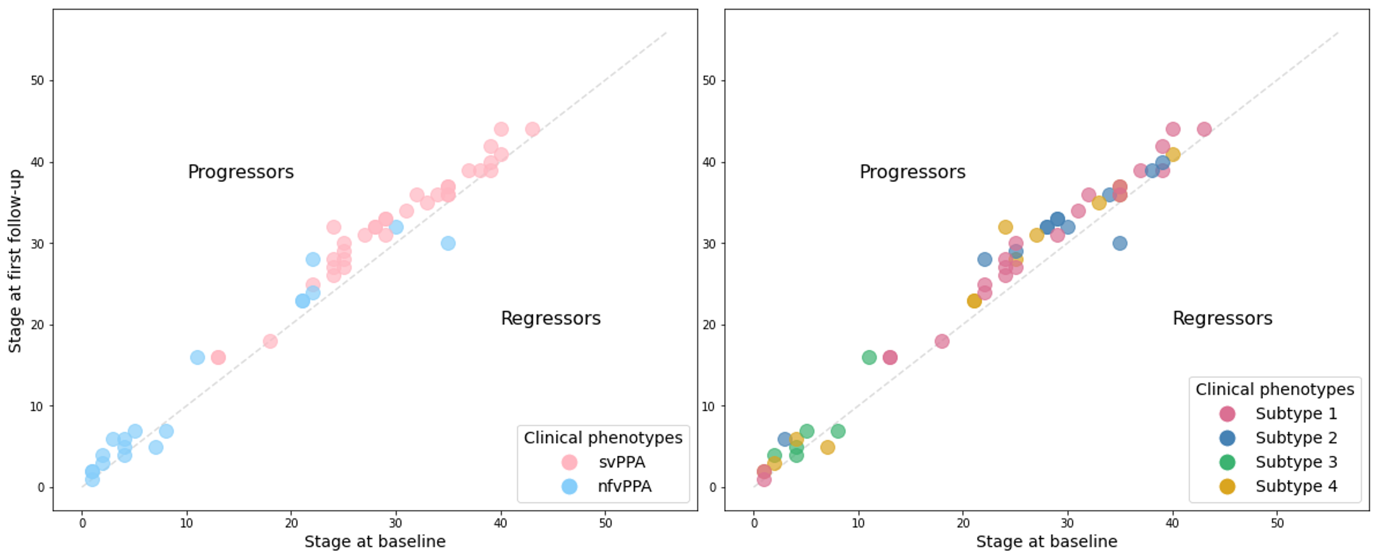

Supplementary Figure 17: **Data-driven stage assignment at baseline and first follow up in the ALLFTD test set.** In the figure on the left patients are colour coded according to clinical diagnosis, and on the right they are colour coded according to subtype. Those above the diagonal progressed at follow up, whilst those below regressed.

Abbreviations: svPPA – semantic variant PPA, nfvPPA – nonfluent/agrammatic variant.

### Post 2010 Dataset

The diagnostic term of lvPPA was enshrined in consensus criteria in 2011.^7,8^ Despite the re-diagnosis of pre-2010 cases by a senior neurologist we were cautious that they were more susceptible to misdiagnosis. To investigate this we re-ran the analysis on a subset of the data, restricted to those individuals diagnosed at Queen Square in or after 2011. The demographics of the post 2010 subset is given in Supplementary Table 6.

#### Post 2010 demographics

|  | | PPA | svPPA | nfvPPA | lvPPA | PPA-nos | Controls |
| --- | --- | --- | --- | --- | --- | --- | --- |
|  | | Queen Square discovery dataset (post 2010) | | | | | |
| n | | 129 | 37 | 52 | 28 | 12 | 121 |
| Sex | | 60F:69M | 16F:21M | 29F:23M | 10F:18M | 5F:7M | 65F:56M |
| Age at onset, years, mean ± SD | | 62.9±7.9 | 55.3±6.7 | 65.6±8.3 | 63.5±7.6 | 59.9±5.9 | - |
| Age at baseline scan, years, mean ± SD | | 67.2±7.8 | 64.3±6.5 | 69.6± 8.3 | 68.2±7.2 | 63.1±5.8 | 61.7± 11.1 |
| Primary pathology, n | |  | | | | | |
|  | Alzheimer’s disease | 2 | 0 | 0 | 2 | 0 | - |
|  | FTLD-tau | 6 | 0 | 4 | 0 | 2 | - |
|  | FTLD-TDP43 | 3 | 1 | 2 | 0 | 0 | - |
| Secondary clinical diagnosis, n | |  | | | | | |
|  | PD | 0 | 0 | 0 | 0 | 0 | - |
|  | PSP | 3 | 0 | 3 | 0 | 0 | - |
|  | CBS | 5 | 0 | 4 | 0 | 1 | - |
|  | PSP/CBS | 1 | 0 | 1 | 0 | 0 | - |
|  | MND | 0 | 0 | 0 | 0 | 0 | - |

Supplementary Table 6: **Post 2010 cohort demographics.** Colours are used to distinguish the three phenotypes: svPPA (pale pink), nfvPPA (pale blue), lvPPA (dark green). Abbreviations: PPA – Primary Progressive Aphasia, svPPA – semantic variant PPA, nfvPPA – nonfluent/agrammatic variant PPA, lvPPA – logopenic variant PPA, PPA-nos – PPA not otherwise specified, FTLD-tau – Frontotemporal lobal degeneration tau, FTLD-TDP43 – frontotemporal lobar degeneration TAR DNA-binding protein 43, PD – Parkinson’s disease, PSP – progressive supranuclear palsy, CBS – corticobasal syndrome, MND – motor neurone disease.

#### Post 2010 results

We used the same ROIs, and same parameters as for the discovery dataset model. Model cross-validation found 2 subtypes to be optimal in this subset. When we forced the model subtype hyperparameter to be four, the four subtypes were similar to those in the discovery cohort.

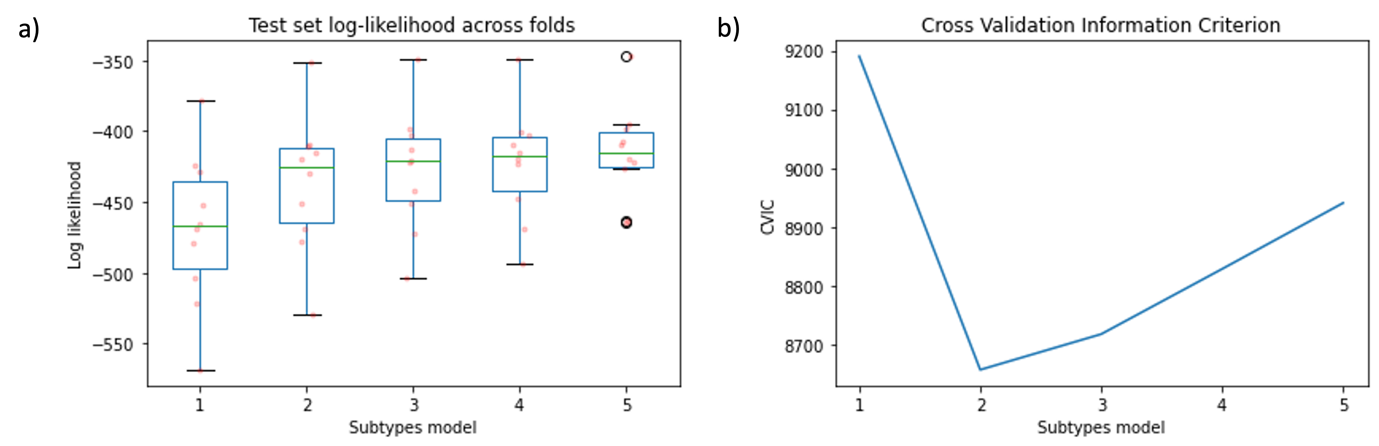

Supplementary Figure 18: a) Test set log likelihood across folds (post 2010 subset) b) Cross validation information criterion for n subtype models (post 2010 subset).

##### Two subtype model

In the two subtype model the first subtype (S1 - left precentral gyrus) there was initial marked atrophy in the left precentral gyrus, followed by parietal lobe and frontal lobe regions. This was followed by diffuse atrophy of the temporal lobe and insula cortex.

In the second subtype (S2 – left insula) there was initially rapid progression through the first and second w-scores in the left insula. This was followed by rapid brain volume change in the left amygdala and the left inferior temporal cortex. There was relative sparing of the frontal lobe and parietal lobe.

In the two subtype model 127/129 patients were assigned above stage 0, and hence deemed to be subtypable, the demographics by subtype are given in Supplementary Table 7. Supplementary Figure 20 compared clinical phenotypes of those assigned to the two subtypes; S2 (left insula) was largely associated with svPPA, whilst S1 (left precentral gyrus) was a mix of nfvPPA and lvPPA.

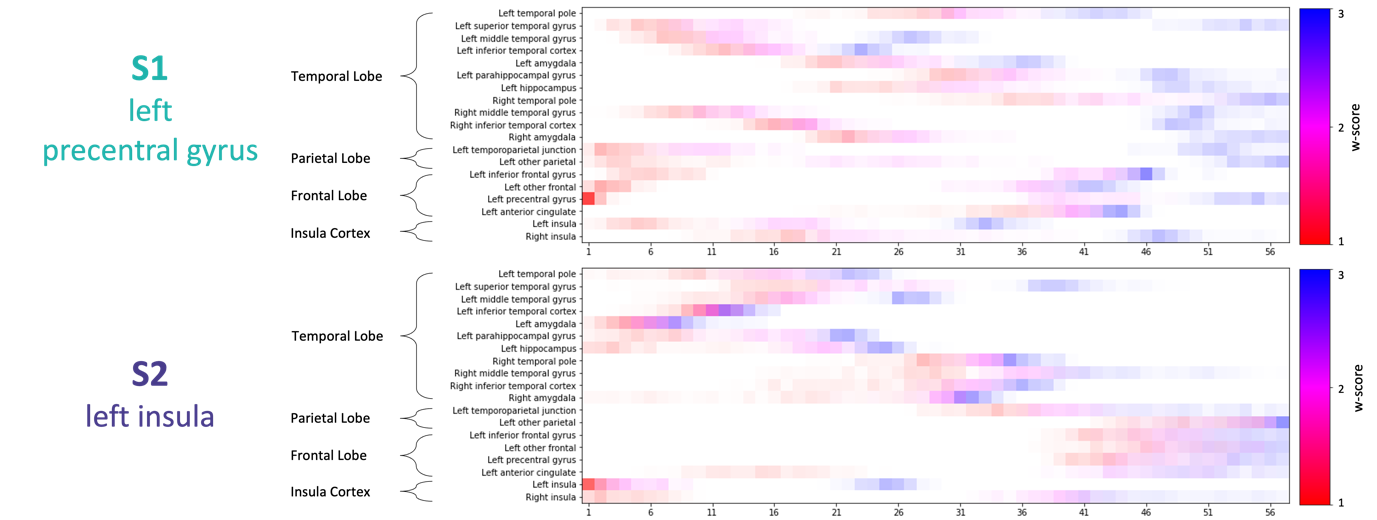

Supplementary Figure 19: **Positional variance diagrams for the two data-driven subtypes in the post 2010 subset.** Along the y-axis are the regions of interest used in the model, grouped by location in the brain. The data-driven stages correspond to the sequence that brain regions become abnormal, with colour representing degree of abnormality (w-score 1: red, w-score 2: pink, w-score 3: blue), and colour density representing model certainty.

|  | | S1 (left precentral gyrus) | S2 (left insula) |
| --- | --- | --- | --- |
|  | | Queen Square discovery dataset (post 2010) | |
| n | | 77 | 50 |
| Sex | | 35F:42M | 23F:27M |
| Age at onset, years, mean ± SD | | 63.5±8.0 | 61.5±7.3 |
| Age at baseline scan, years, mean ± SD | | 68.1±8.0 | 65.6±6.9 |
| Diagnosis, n | |  | |
|  | svPPA | 6 | 31 |
|  | nfvPPA | 42 | 8 |
|  | lvPPA | 24 | 4 |
|  | PPA-nos | 5 | 7 |
| Primary pathology, n | |  | |
|  | Alzheimer’s disease | 2 | 0 |
|  | FTLD-tau | 3 | 3 |
|  | FTLD-TDP43 type C | 1 | 2 |
| Secondary diagnosis, n | |  | |
|  | PD | 0 | 0 |
|  | PSP | 3 | 0 |
|  | CBS | 3 | 2 |
|  | PSP/CBS | 1 | 0 |
|  | MND | 0 | 0 |

Supplementary Table 7: **Post 2010 subtype demographics in two subtype model.** Colours are used to distinguish the four data driven subtypes: S1 (pink), S2 (blue), S3 (green), S4 (yellow). Abbreviations: PPA – Primary Progressive Aphasia, svPPA – semantic variant PPA, nfvPPA – nonfluent/agrammatic variant PPA, lvPPA – logopenic variant PPA, PPA-nos – PPA not otherwise specified, FTLD-tau – Frontotemporal lobal degeneration tau, FTLD-TDP43 – frontotemporal lobar degeneration TAR DNA-binding protein 43, PD – Parkinson’s disease, PSP – progressive supranuclear palsy, CBS – corticobasal syndrome, MND – motor neurone disease.

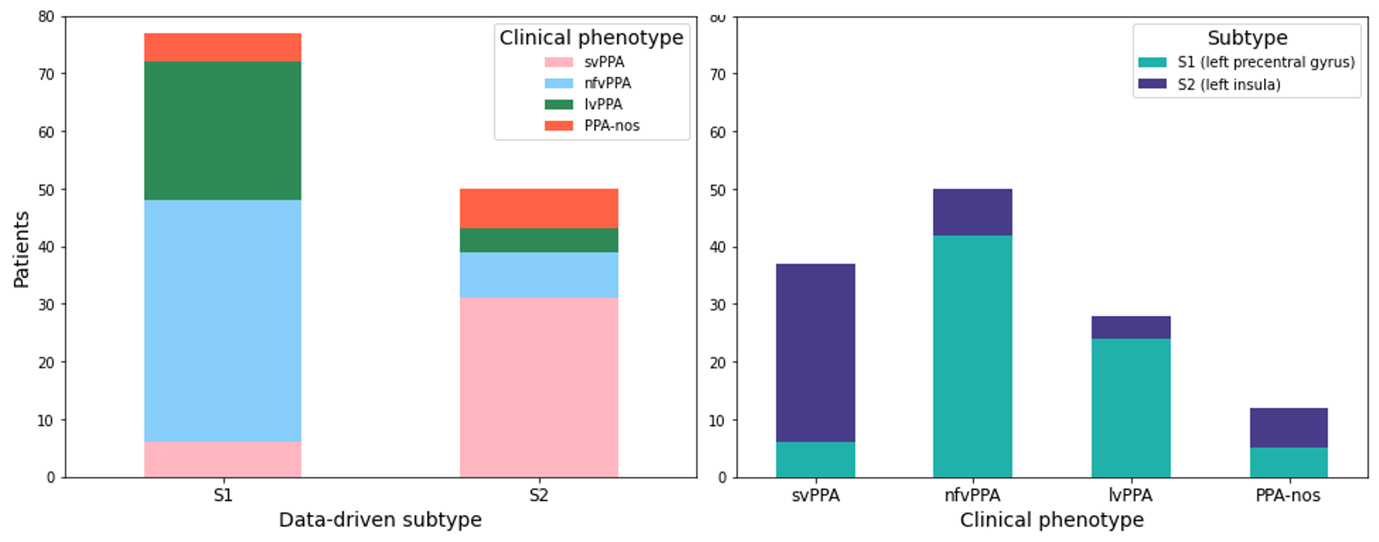

Supplementary Figure 20: **Post 2010 2 subtype model - comparison between data-driven subtype assignment and clinical diagnosis.** The stacked bar chart on the left shows the number of patients with each clinical diagnosis by data-driven subtype assignment. The figure on the right shows the number of patients who were assigned each data-driven subtype by clinical diagnosis.

Abbreviations: svPPA – semantic variant PPA, nfvPPA – nonfluent/agrammatic variant PPA, lvPPA – logopenic variant PPA, PPA-nos – PPA not otherwise specified.

##### Four subtype model

In the four subtype model the positional variance diagrams were largely similar to those seen in the full Queen Square discovery dataset. Visually compared to the positional variance diagrams for the full Queen Square dataset, the four subtypes had higher positional variance, likely due to the lower sample size.

In the four subtype model 127/129 patients were assigned above stage 0, and hence deemed to be subtypable, their demographics are given in Supplementary Table 8. Supplementary Figure 22 compared clinical phenotypes of those assigned to the four subtypes; similarly to the full dataset analysis there was a clear correspondence between S1 and svPPA, with more mixed association between S2/S3/S4 and lvPPA and nfvPPA.

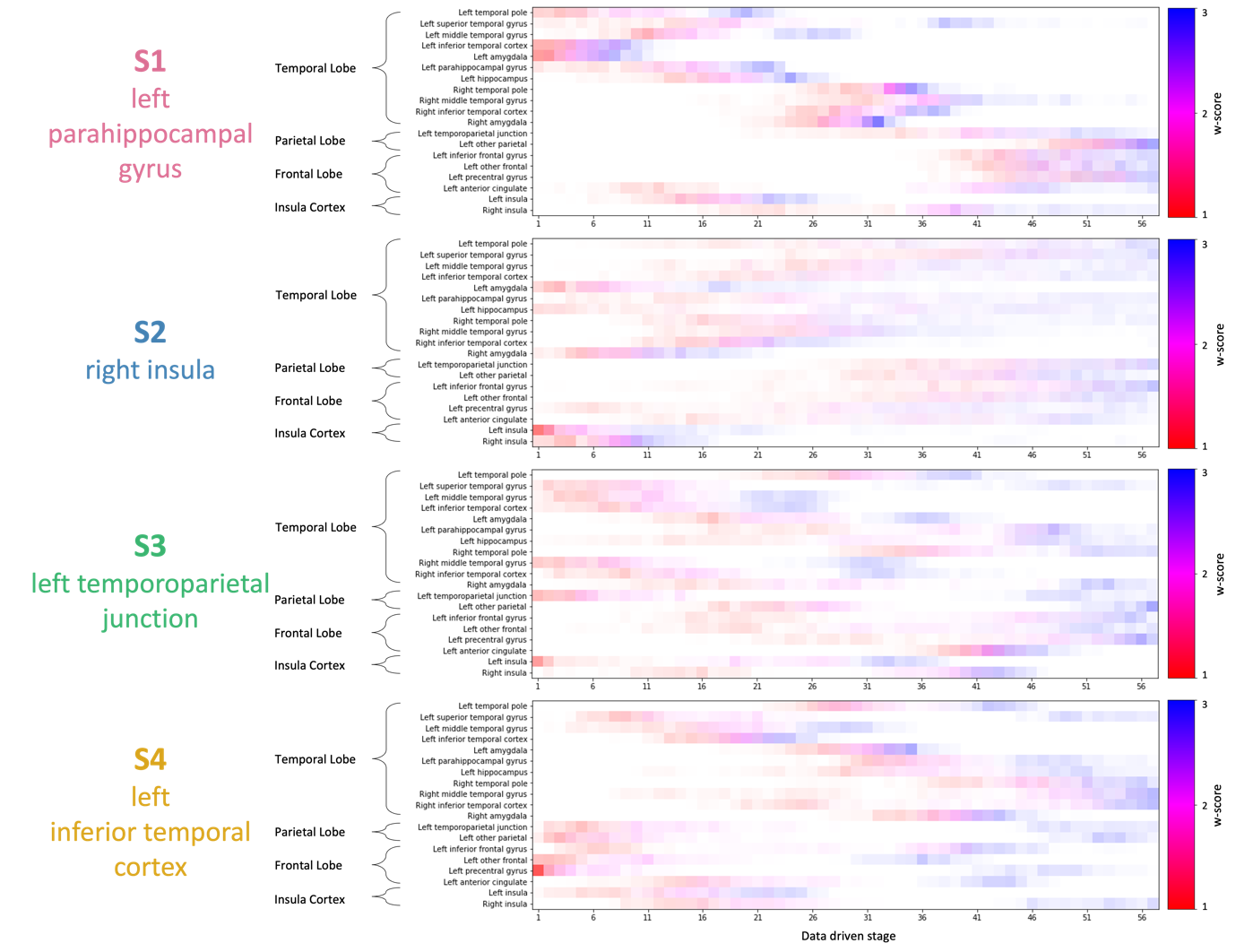

Supplementary Figure 21: **Positional variance diagrams for the four data-driven subtypes in the post 2010 subset.** Along the y-axis are the regions of interest used in the model, grouped by location in the brain. The data-driven stages correspond to the sequence that brain regions become abnormal, with colour representing degree of abnormality (w-score 1: red, w-score 2: pink, w-score 3: blue), and colour density representing model certainty.

|  | | S1 (left parahippocampal gyrus) | S2 (right insula) | S3 (left temporoparietal junction) | S4 (left inferior frontal cortex) |
| --- | --- | --- | --- | --- | --- |
|  | | Queen Square discovery dataset (post 2010) | | | |
| n | | 42 | 9 | 42 | 34 |
| Sex | | 19F:23M | 4F:5M | 20F:22M | 15F:19M |
| Age at onset, years, mean ± SD | | 60.8±7.0 | 64.8±7.0 | 60.8±7.2 | 67.1±8.0 |
| Age at baseline scan, years, mean ± SD | | 64.9±6.5 | 68.3±7.3 | 65.1±6.8 | 71.9±8.2 |
| Diagnosis, n | |  | | | |
|  | svPPA | 30 | 1 | 3 | 3 |
|  | nfvPPA | 5 | 6 | 15 | 24 |
|  | lvPPA | 1 | 1 | 22 | 4 |
|  | PPA-nos | 6 | 1 | 2 | 3 |
| Primary pathology, n | |  |  |  |  |
|  | Alzheimer’s disease | 0 | 0 | 2 | 0 |
|  | FTLD-tau | 2 | 1 | 0 | 3 |
|  | FTLD-TDP43 type C | 2 | 0 | 0 | 1 |
| Secondary diagnosis, n | |  |  |  |  |
|  | PD | 0 | 0 | 0 | 0 |
|  | PSP | 0 | 1 | 1 | 1 |
|  | CBS | 1 | 1 | 0 | 2 |
|  | PSP/CBS | 0 | 0 | 0 | 1 |
|  | MND | 0 | 0 | 0 | 0 |

Supplementary Table 8: **Post 2010 subtype demographics in four subtype model.** Colours are used to distinguish the four data driven subtypes: S1 (pink), S2 (blue), S3 (green), S4 (yellow). Abbreviations: PPA – Primary Progressive Aphasia, svPPA – semantic variant PPA, nfvPPA – nonfluent/agrammatic variant PPA, lvPPA – logopenic variant PPA, PPA-nos – PPA not otherwise specified, FTLD-tau – Frontotemporal lobal degeneration tau, FTLD-TDP43 – frontotemporal lobar degeneration TAR DNA-binding protein 43, PD – Parkinson’s disease, PSP – progressive supranuclear palsy, CBS – corticobasal syndrome, MND – motor neurone disease.

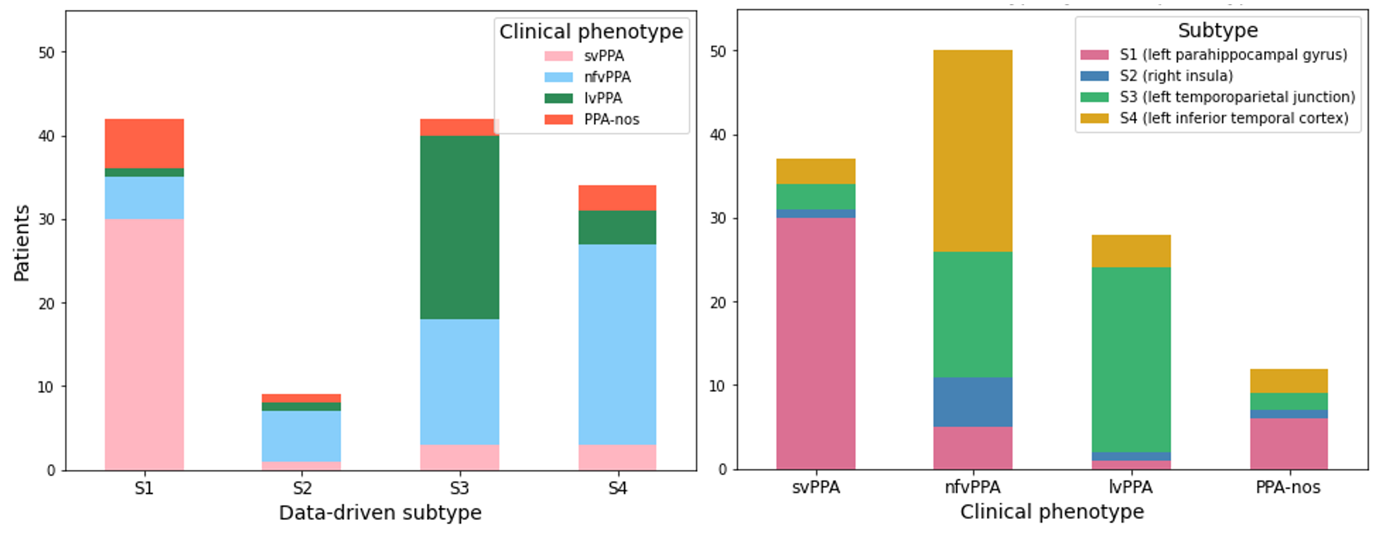

Supplementary Figure 22: **Post 2010 four subtype model - comparison between data-driven subtype assignment and clinical diagnosis.** The stacked bar chart on the left shows the number of patients with each clinical diagnosis by data-driven subtype assignment. The figure on the right shows the number of patients who were assigned each data-driven subtype by clinical diagnosis.

Abbreviations: svPPA – semantic variant PPA, nfvPPA – nonfluent/agrammatic variant PPA, lvPPA – logopenic variant PPA, PPA-nos – PPA not otherwise specified.

#### Post-2010 summary

This reanalysis aimed to investigate how the re-diagnosis of earlier cases might have influenced the outcomes. Cross-validation of the model indicated that two subtypes were optimal in the post-2010 dataset — likely due to the considerably reduced sample size of the post-2010 dataset (n=129 of 270), and subset class imbalance across diagnoses (40.3% of the subset were diagnosed with nfvPPA), compared to the full dataset. However, the four-subtype model in the post-2010 dataset closely resembled that of the entire discovery dataset, supporting the stability of the four subtype in the Queen Square discovery dataset.
